# Study report CAMOPED study

**DOI:** 10.1101/2024.01.25.24301714

**Authors:** A. Schraplau, W. Petersen, M. Herbort, B. Lischke, J. Höher, R. Becker, N. Streich, U. Stöckle, C. Schmidt-Lucke

## Abstract

The following report was prepared as part of a trial guideline in accordance with Section 137e (1) SGB V to evaluate the benefits of the use of CAM splints for self-application by patients in the post-surgical rehabilitative treatment of ruptures of the anterior cruciate ligament.

The primary aim of the study project was to test the hypothesis that rehabilitation with CAMOped in addition to standardised rehabilitation (intervention group, IG) is superior to standardised rehabilitation alone (control group, KG) in terms of knee joint function measured with the sIKDC (subjective assessment) at 6 weeks post surgery (FU2) compared to the time of assessment immediately post-surgical (T4).

Between June 2020 and January 2023, 105 patients (m:w 62:43, age 33 ± 11 years) were randomised in a stratified manner (50 IG and 55 KG) and included in the study, which was evaluated using an intention-to-treat (ITT) approach. 88 subjects had a simple anterior cruciate ligament rupture, 17 subjects also received a meniscal intervention. The study was discontinued for ethical reasons when the CPM splint was included in standard care and thus not carried out in accordance with the protocol for the CG in January 2023.

There were 87 usable data sets (42 IG and 45 KG) for analysing the primary research question. With regard to the relevant preoperative influencing variables (age, Tegner score, injury, sex ratio or oIKDC) at the time of study inclusion (T0), both study groups were equal. In the IG, 5 patients received an additional meniscus intervention (10% of the IG), while in the KG the proportion was 22% with 12 patients.

The CAM splint was used for a period of 3 weeks preoperatively and 6 [5-7] weeks post-surgical.

At post surgery discharge (time point T4), knee joint function (sIKDC) was significantly higher (higher knee joint function) in the IG compared to the KG (n=87, 33.0 [23.0 - 46.0] vs. 28.0 [21.0 - 33.0], p=0.024). Three weeks post-surgical (FU1, n=85), knee joint function was the same in both groups. At the time of the follow-up examinations after 6 weeks (FU2, time of recording the primary endpoint), there was no difference in the sIKDC score between the intervention and control groups (n=87, 56.5 [48.0 - 64.0] vs. 54.0 [48.0 - 63.0], p=n.s.).

The prespecified analysis, in which the change at the follow-up time after 6 weeks post-surgical (FU2) was analysed in relation to the assessment at post surgery discharge (T4), showed that the absolute change in knee joint function was significantly lower in the IG (n= 42) than in the KG (24.5 [12.0 - 31.0] vs. 29.0 [18.0 - 35.0], p=0.041). After 12 weeks (FU3), the subjective knee joint function was identical in both treatment groups. During the one-year follow-up period, 91 adverse events occurred, including 8 serious adverse events, each unrelated to the investigational product.

From 12/2021 - the date on which the CPM splint was included in standard care - the high number of refusals to participate in a study without CPM splints in the control group by patients and the number of protocol violations and dropouts was striking. After conducting the interim analysis on 19 September 2022 and subsequent discussion with the LKP and the principal investigators, this led to the principal investigator discontinuing the study for ethical reasons. A respective letter in german from the PI to the sponsor of the study can be requested from the study contact.

## I Introduction

### 2. Background / Objectives

#### 2.1. Scientific background and rationale of the study

The CAMOped, an active movement splint, has been used since 2000 for rehabilitation after hip or knee injuries with the aim of improving mobility and functional stability and shortening the rehabilitation phase.

The benefits of the use of active knee motion splints (CAM splints) for self-application at home by patients in the treatment of ruptures of the anterior cruciate ligament had not yet been sufficiently proven, but the method had the potential to be a necessary treatment alternative in accordance with the decision of the Federal Joint Committee (G-BA) of 5 September 2019. There were no known ongoing studies whose results could be used to close the knowledge gap. At its meeting on 16 August 2018, the G-BA therefore decided to suspend the procedure in accordance with Section 135 (1) sentence 1 SGB V to assess the use of CAM splints for self-application by patients in the treatment of ruptures of the anterior cruciate ligament and to begin consultations on a trial guideline in accordance with Section 137e (1) SGB V in order to gain the necessary knowledge for a final assessment of the benefits.

The CPM splint and CAM splint are used after reconstruction of the ACLR to alleviate the negative effects of immobilisation. Both splints can be used immediately in the post-operative phase and the flexion of the knee is adapted to the current findings of the knee. The aims of the CPM splint are to increase the range of motion, reduce swelling, reduce pain, prevent capsular contractures and improve the orientation and strength of the collagen fibres in the ACL graft, thereby supporting the overall healing process.^1 2^ The CAM splint, in which the patient performs an active movement using the non-operated leg, has the advantage over the CPM splint of specifically stimulating the proprioceptors. Neuromuscular coupling is maintained by recalling physiological walking and turning patterns.^3^ Active movement with the CAM splint specifically stimulates the proprioceptors by placing high demands on the coordination of knee joint movement.^4^ Active movement also reduces muscle atrophy.^5^ Friemert concludes that the use of a CAM splint in the immediate post-surgical phase significantly improves the proprioceptive deficit compared to physiotherapy alone.^6^

In contrast to knee orthoses, CAM splints are pedalling machines in which both legs are involved during the movement exercise and only guided movements are possible. As the foot is fixed in the CAM splint, the treatment follows the closed chain principle. According to the manufacturer, the sequence of movements prescribed by the medical device enables early, active training at home.^7^ According to the manufacturer of the CAM splints, their use is recommended from the 4th post-surgical day (see **Error! Reference source not found**.). The CAM splints are intended to promote early proprioception, which is impaired by the ACL rupture, and thus restore functional stability (Available in German on request).

IQWiG conducted a literature search up to 17 January 2017 with the aim of assessing the benefits of (self-)use of active knee braces after conservative and surgical treatment in patients with anterior cruciate ligament rupture with regard to patient-relevant outcomes. The report N16-01 was published on 11 May 2017.^8^

In only 2 publications^6 9^ data were available from clinics, but not from the home setting. Both studies tested the CAM splints post-surgical and included 50 ^9^and 60^6^ participants. The smaller study compared the CAM splint against a power-operated passive splint (CPM)^6^ while the larger study tested the CAM splint against follow-up treatment (physiotherapy).^9^

Both studies have shortcomings in their implementation, such as unclear allocation of participants to the treatment groups (dubiously conducted randomisation), continuous recruitment, 90% male participants (in contrast to the gender distribution described in the literature), lack of recording of pain medication intake, short follow-up period (7 days), lack of long-term observation.

Predefined endpoints were range of motion (PROM) and pain. PROM cannot be analysed in one study ^9^ as there was already a result-distorting difference between the two groups preoperatively. CAM and CPM appeared to have the same effect on PROM. The recording of pain is not clear. The authors of the report doubt that both studies were able to recognise the relevant target criterion, depth sensitivity, as in their view it is not ensured “that the test used for data collection really captures depth sensitivity.”

To evaluate the benefit of the use of CAM splints for self-application by patients in the rehabilitation phase after surgical treatment of ruptures of the anterior cruciate ligament as part of a trial guideline in accordance with § 137e paragraph 1 SGB V for evaluation, the evaluation was carried out until the clinical trial was discontinued as required by the LKP.

The test product is an active motion splint (CAM) with the trade name CAMOped for use in the home by patients in accordance with technical instructions / instructions for use (see Appendix **Error! Reference source not found**.).

Recommended frequency / duration of use: 4 x daily for 20 minutes until knee flexion of approx. 110° is achieved (estimated treatment duration 3 to 6 weeks)

Certified for the following indications:

1. cruciate ligament ruptures
2. repositioning osteotomies
3. knee and hip prostheses
4. reconstructive procedures on menisci and articular cartilage
5. after hip/knee ASK in combination with synovectomy
6. retropatellar damage
7. ankle fracture

#### 2.2. Research question / hypothesis

The aim of this study was therefore to enable the evaluation of the benefits of this method at a level of knowledge that is sufficiently reliable for subsequent guideline decisions. The primary aim of the study project was to test the hypothesis as to whether, following surgical reconstruction of a unilateral rupture of the anterior cruciate ligament (ACL), rehabilitation with CAMOped in addition to standardised rehabilitation alone is superior to standardised rehabilitation alone after 6 weeks post-surgical in terms of knee joint function as measured by the IKDC (subjective assessment).

##### Primary hypothesis: CAMOped leads to an improvement in joint function

Test: Comparison of the relative change in knee joint function to the initial findings between the two treatment groups using the IKDC 2000 (patient questionnaire for subjective assessment of the knee, sIKDC)

**Secondary objectives** were to demonstrate (Available in German on request)

- the safety of the test product,
- the improvement of quality of life (KOOS, Q1 to Q4),
- the acceleration of rehabilitation,
- influencing proprioception with a lower re-injury rate,
- optimisation of measurement accuracy in the early rehabilitation phase through more suitable measurement instruments (GAS, modified IKDC (now used under the name LERAS-13),
- the economic efficiency of CAMOped,
- the comparability of different surgical procedures (graft selection),
- the superiority of sonographic findings over palpation,
- the non-inferiority of the CAM splint compared to the CPM splint in the subgroup of ACLR with concomitant injuries requiring intervention (subgroup).

## II Methodology

### 3. Study design

A multicentre, randomised, controlled and single-blinded study was conducted to prove the superiority of an active movement splint for self-application at home in the treatment of ruptures of the anterior cruciate ligament in addition to standardised physiotherapeutic rehabilitation.

The control group in the study was defined in agreement with the G-BA according to standard guideline practice without the use of CAMOped. This was ethically justifiable at the start of the study in 2019.

In 5 study centres in Germany, patients were divided into two treatment arms and followed up for a period of 2 years.

The study design was not changed.

### 4. Patients

Potential study candidates were consecutively approached about their possible participation in the study according to the inclusion and exclusion criteria.

A patient was considered included after consent documented by signature and randomised assignment to a treatment arm. This point in time marked the start of the study-related procedures, documentation and recording/follow-up of adverse events.

The selection of inclusion and exclusion criteria was chosen in such a way that a broad patient collective with unilateral ACLR in Germany is represented.

To ensure transferability to the target population, all suitable patients were included in the study based on the inclusion and exclusion criteria. Only patients for whom a surgical procedure was primarily planned were recruited for this study.

Based on clinical experience, it was to be assumed that patients with primary ACLR without injury to the other ligaments and different injuries to the menisci (approx. 30% of the population) were more likely to be affected ^*10 11*^. In the latter, it was expected that a meniscus refixation and at most a partial resection would be necessary. Patients who receive such an intervention must be analysed separately (separate subgroup) and patients should be recruited until the estimated number of cases is reached for sufficient significance for patients after ACL reconstruction without meniscus damage.

#### 4.1. Inclusion and exclusion criteria

Initially, it was decided that only people with isolated ACLR should be included. After intraoperative intervention decisions had to be made for concomitant injuries requiring intervention (in particular meniscus lesions) with the guideline-compliant use of post surgery CPM splints in rehabilitation, a subgroup was formed for these patients on 14 December 2020 after consultation with the G-BA (Available in German on request) in order to also include this common indication in the review.

The inclusion and exclusion criteria of the study were:

#### 4.2. Environment / place of realisation

Patients for whom surgical treatment of the ACLR was planned were included. The interventional procedure was planned according to the assessment of the treating surgeon. For the planning of the clinically indicated procedure, the assessment of general condition, sporting and/or physical activity, MRI findings and concomitant injuries was routinely carried out in accordance with current guidelines ^12^. The operation was performed on an outpatient or inpatient basis in accordance with the site and the standard procedure of the respective centres.

In this multicentre, Germany-wide study, patients were included in the following trial centres:

- Martin Luther Hospital Berlin
- Sportsclinic Cologne Cologne
- Orthopaedic Surgery Munich
- University Hospital Brandenburg an der Havel
- Sportopaedie Heidelberg

### 5. Treatment / Intervention

The operations could be performed on an outpatient or inpatient basis. Depending on the centre, femoral catheters or saphenous blocks were used for analgesia. Post-operative pain therapy was carried out according to the symptoms with NSAIDs, tramadol, metamizole or similar if required. Intra-articular drains (10 mm) are usually removed after one day.

Post-surgical, all patients received standardised rehabilitation treatment in accordance with the current procedure and reimbursement by the statutory health insurance funds of 12 treatment units (usually 2 consecutive prescriptions for 6 x manual and physiotherapeutic therapy) for dynamic stabilisation of the knee joint (Available in German on request).

Standardised rehabilitation was carried out for this study in order to be able to compare the centres with each other. For standardisation purposes, all patients (IG and KG) completed a form containing the treatment recommendations of the study physicians in strict accordance with the tabular list in the work by Niederer, et al. (2019) (Available in German on request.).^13^

These physiotherapy sheets were given to the patient to present to the physiotherapy practice. For their part, the physiotherapists documented weekly specifications on the content and exercise times of the patients’ own exercise programmes. These documentation forms were brought to each follow-up visit, photocopied and filed in the respective patient’s study folder and continuously updated for the next episode. In addition, the patients documented the exercises they actually performed in an electronic diary.

This approach made it possible to record how the recommended guidelines were implemented and to establish comparability between the two treatment arms.

The physiotherapy was carried out in physiotherapy practices chosen by the patient. All 5 principal investigators receive referrals from a larger radius, which is spread across the city itself and also from other districts of the capital. It is contrary to the clinical and care reality in Germany to send patients to physiotherapy practices that may be far away for a standardised rehabilitation measure. Outside of this study and after completion of the study, patients are also treated in different practices that carry out rehabilitation according to their own standards.

In order to record the qualifications of physiotherapy treatment, all physiotherapy practices are contacted and surveyed electronically and documented with regard to the following aspects:

1. Frequency of treatment of ACLR in practice
2. Number of years of certification of the treating physiotherapist
3. Standard therapy for treatment after ACLR in the practice
4. Decision scheme for the rehabilitation programme applied in the case of postintervention ACLR

#### 5.1. Subgroup of patients with concomitant knee joint injuries requiring intervention diagnosed intraoperatively

A so-called “restrictive rehabilitation scheme” was used for patients who, despite careful planning of the operation, still had knee findings requiring intervention intraoperatively (Available in German on request) and who had to be treated unilaterally in accordance with the guidelines:

Factors that indicate a so-called “restrictive rehabilitation programme” include, for example, the need for a meniscus suture or more pronounced other concomitant injuries in ACLR. At some centres, treatment with CPM splints in addition to physiotherapy was recommended as standard for patients with concomitant knee injuries requiring intraoperative intervention and it was considered unethical to withhold a CPM splint from these patients.

Thus, rehabilitation for patients with concomitant injury requiring intervention who were randomised preoperatively to the control group was carried out with a CPM splint. Patients randomised to the CAMOped group remained in this group.

#### 5.2. Adherence to therapy / issue and return of test products

After randomisation of a patient by an unblinded MEDIACC employee, the latter sent an order form to the manufacturer stating the patient’s name, the delivery address and the desired delivery date as well as the respective principal investigator/trial centre as prescriber.

This information on the form was transmitted electronically to the regional OPED employees in accordance with the manufacturer’s existing standard instructions in a standardised form that did not indicate that the patient was a study patient. Delivery was then carried out by a delivery service to the patient or by an OPED referrer. In most cases, the patient was personally instructed before the first use. In exceptional cases, due to physical distance, legal requirements or the patient’s wishes, this could also be done by telephone and supported by digital media. The patient acknowledged receipt of the test product to the OPED employee or - in the case of instruction by telephone - the patient sent the confirmation to OPED with a prepaid envelope.

After treatment or in the event of a complaint, the patient returned CAMOped. The pre-paid return slip and the information were already enclosed on delivery. There was no further contact between the manufacturer and the study patients, except for familiarisation or to solve documented technical problems. In particular, there was to be no contact with the study patients on the manufacturer’s initiative. This was regularly checked with the patients.

In addition to the standard physiotherapy treatment, patients in the intervention group were to use CAMOped independently in their own home environment for 4 x 20 minutes a day.

After recruitment and inclusion according to the inclusion and exclusion criteria (see 4.1), the patients were randomly assigned to the treatment arms according to the stratification characteristics (age and physical activity according to the Tegner score):

- Control group: standard physiotherapy
- Intervention group: standard physiotherapy and use of the CAMOped

Patients in the group who received CAMOped were instructed in the use of the device by OPED staff at home in accordance with the manufacturer’s standard procedures. The patients were able to use the device pre-operatively.

After the operation, the rehabilitation phase followed with or without CAMOped use (according to randomisation).

In Illustration 1 shows the study process schematically.

**Illustration 1.**
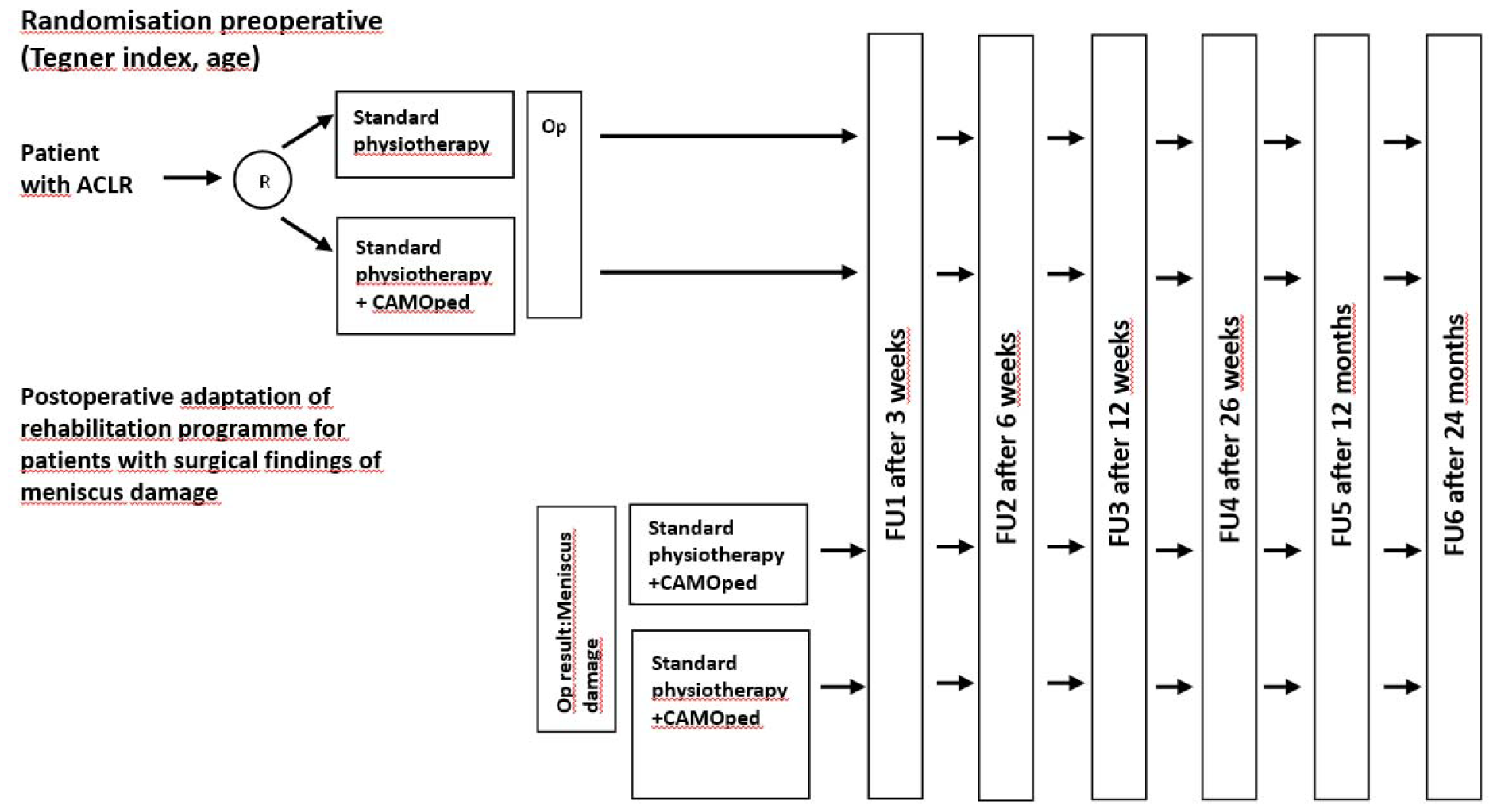
Study programme. Abbreviations: FU, Follow-Up; ACLR: Anterior cruciate ligament rupture ; R: Randomization.

If additional meniscus damage was found during the operation, the patients in the control group also received a CPM splint.

Examinations were planned at T0 (preoperative), T2 (preoperative, if T0 is more than 2 weeks before surgery), T4 (post surgery, at discharge) and at various points in the post surgery follow-up: FU1 (3 weeks), FU2 (6 weeks), FU3 (12 weeks), FU4 (6 months), FU5 (12 months), FU6 (24 months).

#### 5.3. Procedure

- After receiving the positive ethics vote, potential patients were approached
- Adaptation of the study design to the requirements of the G-BA to ensure the suitability of the study concept for the final benefit assessment and to increase the certainty of the results
- Recruitment of patients with inclusion and exclusion criteria for planned surgical procedures
- Informed consent of the patient
- Instruction in CAMOped by OPED staff at home (in accordance with randomization and standard procedures at the manufacturer)
- Use of CAMOped (according to randomization)
- Operation
- Rehabilitation with or without CAMOped (according to randomization)

#### 5.4. Visiting plan

The ward round schedule is shown in Table 2. In addition to the medical history and physical examination, the visits include the completion of questionnaires by the examination staff.

**Table 1:**
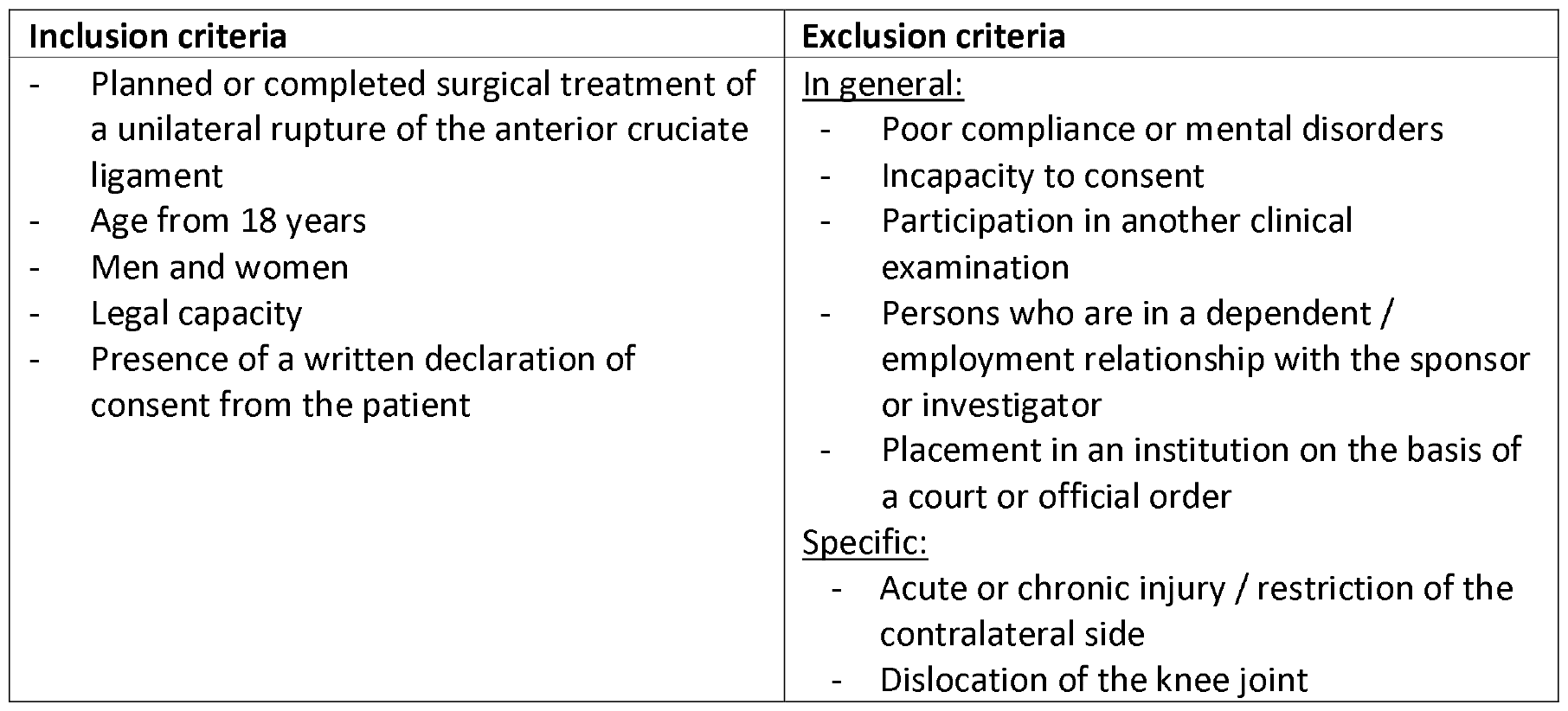
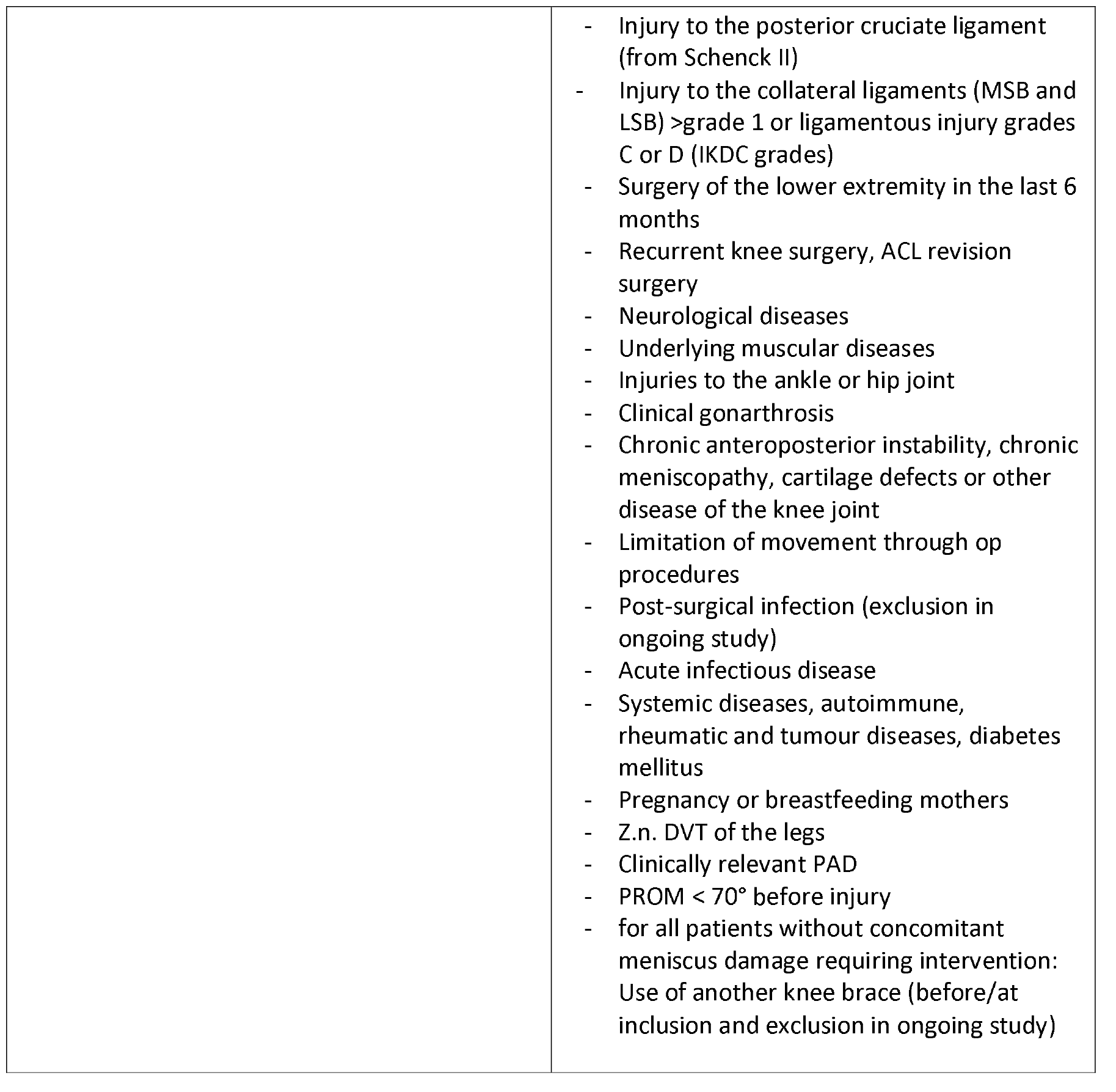
Inclusion and exclusion criteria CAMOPED study.

**Table 2:**
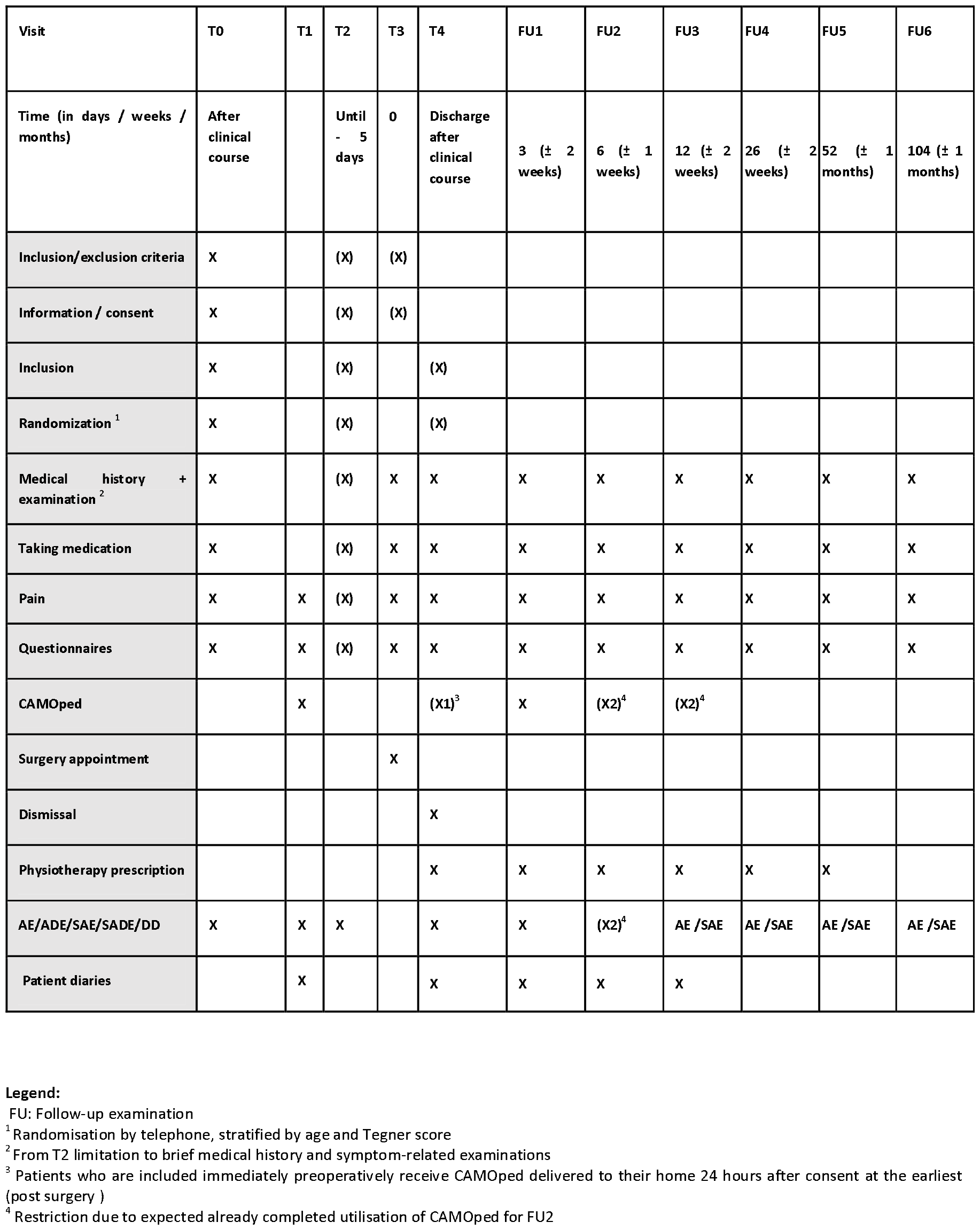
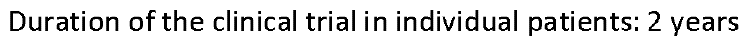
Examinations as part of the clinical study (visit plan)

**Table 3:** One-sided group-sequential experimental design for 4 interim analyses.

| Analysis | N | Z | Nominal p Spend |
| --- | --- | --- | --- |
| 1 | 78 | 2.85 | 0.0022 |
| 2 | 130 | 2.21 | 0.0123 |
| 3 | 156 | 2.01 | 0.0118 |
| 4 | 208 | 1.74 | 0.0237 |

The study participants completed questionnaires electronically via an online platform. They received the requests to complete the questionnaires, including the access links to the corresponding study time points, in advance by email.

Efficacy and safety parameters were recorded via physical examination and questionnaires as described in the ward round schedule. Validated questionnaires were used as well as recording forms in which adverse events were asked about and documented at the time of each visit (see Appendix 1 RCT CAMOped V1.2 trial protocol 20201214, section 4.7.1).

### 6. End points

#### 6.1. Measurement of efficacy and safety parameters

##### Target values

###### Primary target figure

Knee joint function after 6 weeks (FU2) compared to the initial examination (T4) measured using the IKDC 2000 (subjective patient questionnaire on current state of health)

###### Secondary and other targets

- Security
- Frequency and severity of adverse events, particularly product-related
- Benefit/risk assessment
- Benefit assessment, medical necessity
- Frequency of re-ruptures and surgical interventions or revision procedures
- Influence of the pre-operative use of CAMOped
- Pain reduction after within the first 6 weeks
- Change in the intake of pain medication after 1 and 3 weeks
- Effusion regression after 3 and 6 weeks, occurrence up to 24 months
- Primary wound closure after 3 weeks
- functional parameters, recording of active and passive range of motion (PROM), strength and mobility tests, jump tests and test procedures used to assess coordinative abilities such as stance stability (subgroup), IKDC-2000, physical examination, Rolimeter, in each case in a side-by-side comparison (3 mm optimal stability, up to 5 mm suboptimal, > 6 mm instability) ^14^;
- Achievement of targets from the goal attainment scale after 3 and 6 weeks (swelling, pain, fluid gait pattern, getting into a car if necessary, cycling) and up to 24 months
- Combined endpoint of knee joint function after 6 weeks (FU2) compared to baseline (T4 measured on the IKDC 2000 (subjective patient questionnaire on current health status) and quality of life (KOOS, Q1 to Q4)
- Achievement of 120° flexion and lack of swelling after 6 weeks
- Time to return to exercise capacity (daily activities, cycling, sport (training or competition) / ability to return to work) - in each of the relevant subgroups
- Time until start of physiotherapy, start and performance of actual therapy, time until the predefined 12 PT sessions are reached
- Exercise capacity as a function of the pre-injury activity index (Tegner score)
- Time to return to sport / ability to return to work depending on the activity index before injury (Tegner score)
- Reduction of impaired rehabilitation and late complications
- Improvement of the functional stability of the knee joint, better coordination and training of the proprioceptors
- Muscular atrophy of the ipsilateral quadriceps muscle
- Self-confidence (ACL-RSI Knee Score) from 3 weeks
- Economic efficiency in domestic use (economic analysis)
- Frequency and length of use of the CAMOped
- User-friendliness of the CAMOped
- Improvement in knee joint function when using the CAMOped pre-operatively
- (subgroup to be analysed separately): Non-inferiority of CAMOpeds compared to CPM splints
- Validation of modified IKDC sheet
- Validation of sonography as an additional measurement tool for therapy decisions and return to sport/competition

###### Security analysis

Adverse and serious adverse events were documented for all patients and made available to the principal investigator in a timely manner.

##### Measuring instruments, questionnaires and evaluation forms

###### Medical history

– Injury history: symptoms, stiffness, pain, activities of daily living, sport and leisure, impact of the affected knee on quality of life, injury history, previous operations
– Activity index
– Pain
– Amount and type of pain medication (pain diaries)
– Special features (AE, SAE)
– Psychological component for the resumption of (competitive) sport
– Rehabilitation measures
– Only for a subgroup of concomitant knee injuries requiring intervention: use of the CPM splint

###### Investigations

– Knee joint examination (lateral comparison): PROM, Lachmann, laxity, leg axis, patella, dislocation, range of motion, effusion, ligaments, compartment, graft morbidity, joint space, wound healing
– Objectively quantified reduction of the knee joint effusion, measured in cm circumference at the level of the joint space and 10 cm and 20 cm cranial to the joint space
– Muscular atrophy
– Functional tests: hopping on one leg, gait pattern (from approx. FU 3)

###### Apparative

– Rolimeter
– Pedometer with CAMOped
– Knee joint effusion: Sonography

###### Activity parameters

- Time between ACLR and surgery
- Time until resumption of work
- Time until resumption of (competitive) sport
- Time to resumption of competition in patients with a high activity index (Tegner score activity level ≥7)
- Psychological component for resumption of sport

###### Safety / study quality

- Number of adverse events (AE) and serious adverse events (SAE)
- Time to occurrence of AE and SAE
- Obtaining AE and SAE for the use of CAMOped
- Number of dropouts, time of dropout
- Reasons for dropping out, reasons are classified if necessary
- Examiner
- Adherence (cumulative duration of use, number of sessions / day or / week, length of use in weeks) to the use of the CAM or CPM splint and the other randomization rehabilitation measures
- Start and length of use of the CAM or CPM splint and the other randomization rehabilitation measures
- Risk-benefit analysis

The following questionnaires and evaluation forms were used and analysed during the course of the study:

- IKDC Knee 2000
- KOOS questionnaire (KOOS-Q1 to Q4)
- Tegner score
- ACL-RSI
- GAS
- Modified subjective IKDC (LERAS-13)
- Recording of adverse and serious adverse events
- Pain questionnaire
- “Back to work and sport and competition”
- Primary wound healing
- Time of rerupture, re-injury
- Utilisation questionnaire for the test product
- Questionnaire for instructing technician
- Physiotherapy questionnaire (doctors, therapists, patients)
- Visual impression atrophy of the quadriceps muscle
- Exclusion of influence by manufacturer
- Sonographic measurement of knee joint effusion

The **IKDC Knee 2000 is** composed as follows:

- Master data
- Demographic data (page 1)
- Medical history (page 2)
- Previous therapy of the cartilage defect (page 3)
- Previous operations (page 4 -7)
- IKDC 2000 knee examination form (page 8)
- Surgical documentation form (page 9 -11)
- Op acquisition (page 12)
- Form for recording cartilage damage according to ICRS (incl. ICRS functional status)
- Patient questionnaire for subjective assessment of the knee according to IKDC (IKDC Score)
- Patient questionnaire on current state of health according to IKDC (SF-36 score is not used)

The **KOOS questionnaire** is divided into 5 independent subscales: pain, symptoms, activities of daily living, functional ability in sport and leisure and quality of life in connection with the affected knee. The subscales are validated and can each be assessed individually, with each subscale containing questions whose answers are scored on a 5-point Likert scale from 0 to 4. For the KOOS Quality of Life (KOOS QoL) subscale used in this study, the values for the MCID and SCB for patients after ACL reconstruction are 25.9 (MCID) and 35.8 (SCB) respectively. ^15^and the standard values to be aimed for after complete rehabilitation are 86 (male) and 79 (female). ^16^ The state of subjective “well-being” is given in with 63 points.

The **modified subjective IKDC (LERAS-13)** was developed as follows:

**Illustration 2.**
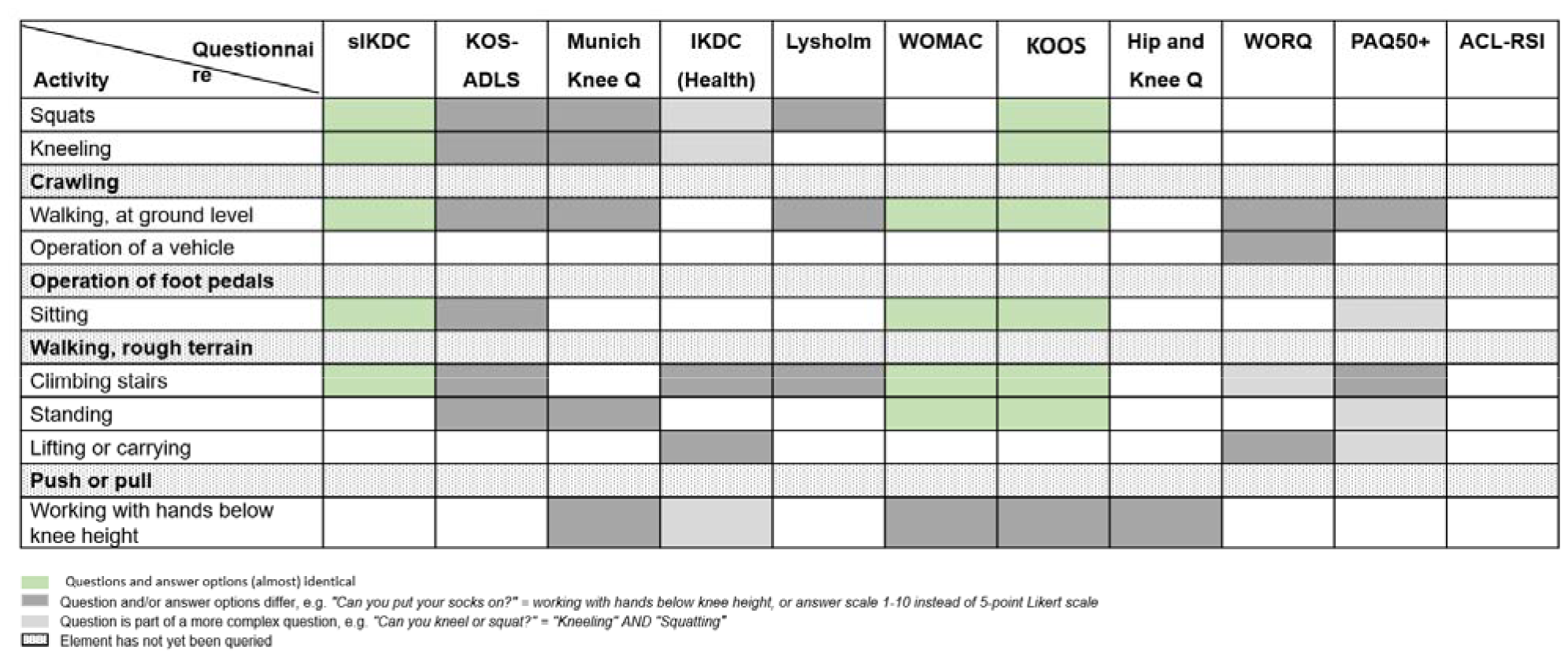
Development of the LERAS-13 using the gap analysis to assess the early rehabilitation phase after lower limb injury.

And contains the following elements:

**Illustration 3.**
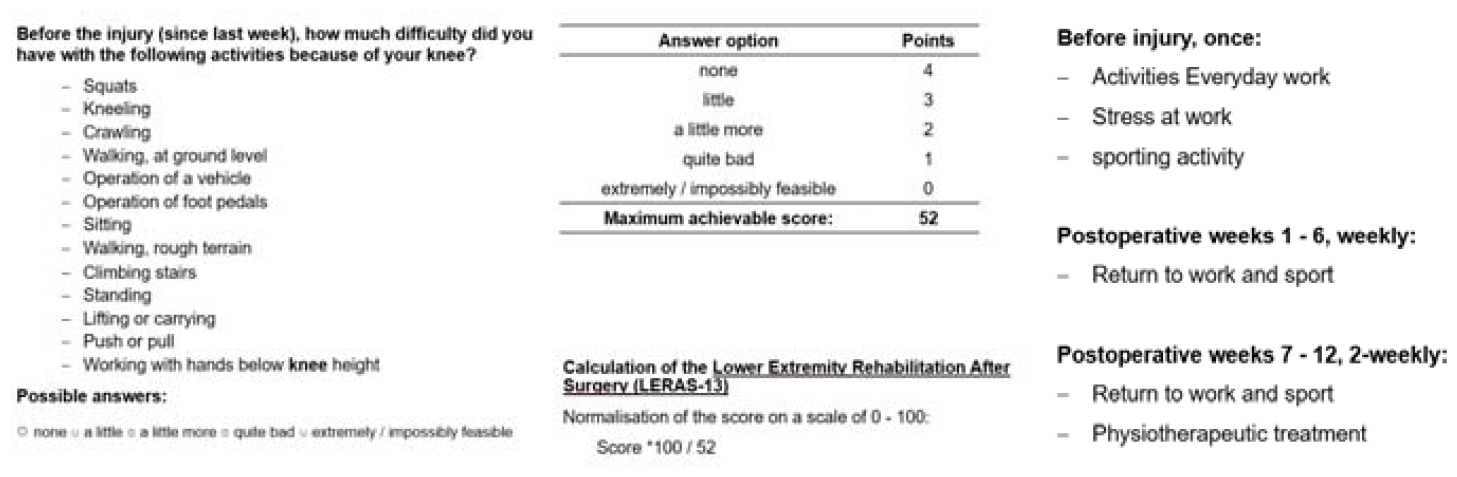
Elements and questions of the LERAS-13

Similar to the subjective IKDC, the number of points given was randomization to the number of answers.

No variables were changed during the study.

### 7. Determination of case numbers

#### 7.1 Determination of case numbers

After the search (August 2020), no data from prospective, randomized, methodologically sound studies were available for a clinically relevant endpoint (as specified on 24 June 2020, the ICDC) on rehabilitation success after surgical treatment of an ACLR, which would have been fully suitable for the case number calculation of the subsequently designed clinical study.

The methodological quality of studies quantifying the success of rehabilitation after ACLR, whether with conservative or surgical treatment, is inadequate and was not suitable for a risk-free estimation of the number of cases for the present study. Only about 12% of the publications adequately describe the interventions (rehabilitation) after ACLR reconstruction. ^18^ There is not a single published, methodologically superior study documenting the treatment success of CPM or CAM splints or other comparable rehabilitative measures after ACL reconstruction using the IKDC or another validated, quantifiable measurement instrument. We have already discussed the problem with the study situation used by IQWIQ elsewhere and hereby refer to it again (our letter dated 22 June 2020).

In particular, the study used in the “Supporting reasons for the decision of the G-BA on the guideline for testing in accordance with Section 137e SGB V “Use of active knee movement splints for self-application at home by patients in the treatment of ruptures of the anterior cruciate ligament” (see p. 11 of the document) by Ingelsrud ^19^ did not seem suitable here. Data from 3 different and late (6, 12 and 24 months) follow-up periods were pooled in the study. The active motion splint allows up to 110° knee flexion and is not usually used in practice after that (see also information sheet from OPED Appendix **Error! Reference source not found**.), but other rehabilitative measures such as cycle ergometers. The influence of CAMOped will then no longer be reliably detectable. It therefore appeared necessary to recalculate the number of cases based on more suitable studies, taking into account early follow-up times (6 weeks). In general, there was also drastic methodological uncertainty (design, disease severity, biometric limitations) regarding the transferability of the effect sizes from these studies to the CAMOPED study.

##### Measuring instrument

The primary endpoint for calculating the number of cases was the intraindividual subjective improvement in joint function 6 weeks post-surgical, measured using the subjective IKDC score.

##### Publications on the calculation of case numbers in the CAMOPED study

In the systematic literature search by 2 independent investigators in PubMed, Cochrane, Prospero and Embase on 10 August 2020, the publications for which the full text and the values of the objective IKDC grades or the subj. IKDC score had to be available, without restriction of language, were found for the estimation of the number of cases for the CAMOPED study with the primary endpoint of superiority on knee joint function using the IKDC measurement tool with the combination of the following keywords:

“ACL AND reconstruction AND rehabilitation AND IKDC”; “ACL AND physiotherapy AND IKDC”;

“acl AND reconstruction AND rehabilitation AND ikdc AND adult” (AWMF and BASE: German keywords)

Languages: en, de, franz, it, span (n=336)

Additional articles identified through other sources (n=13)

After excluding duplicates, the headings of 292 articles were checked for relevance; papers on long-term courses (>1 year), comparison of surgical techniques, adolescents and children, age comparisons, meniscus or multiple ligament damage were excluded. The full texts of the remaining 117 publications were checked for suitability and the full texts of 24 were used for data extraction. Where necessary, further papers were searched for manually.

Data were extracted from 24 studies that were deemed suitable for estimating the success of rehabilitation on knee joint function after ACL reconstruction. Many studies did not contain any information on knee joint function before the injury or preoperatively or immediately post-surgical and were therefore severely impaired or unsuitable in terms of their informative value. The methodological quality of most studies was low with a high risk of bias. Only group comparisons were analysed instead of intra-individual progressions, and in most cases it was not possible to assess with certainty whether complete data sets were compared. However, all studies presented below were of higher quality than those presented in Ingelsrud ^19^ and used to assess the reimbursability of CPM splints.

The referenced works and the extracts of the measured values as well as a brief commentary can be available in German on request.

To calculate the number of cases, it was assumed that the IKDC score (*International Knee Documentation Subjective Knee Form*) was used to measure the primary endpoint six weeks after surgery. The IKDC consists of an objective and a subjective part. The objective part should be answered by a physician and covers the areas of effusion, ligament impairment and functional limitations on an ordinal scale with the gradations “normal”, “near normal”, “abnormal” and “severely abnormal”. Several publications recommend that the objective part should not be used to monitor the course of therapy as, unlike other instruments, it is not sufficiently sensitive to changes in knee joint function (Revenas^20^ Risberg^21^). The subjective part of the IKDC records the changes in symptoms, sports and everyday activities and function as assessed by the patient using rating scales, which are aggregated into a total score. A maximum of 87 points can be achieved; for better interpretation, the value achieved is randomization to 100. The subjective IKDC score was used as the basis for determining the number of cases, whereby the value was determined both before and immediately after the operation and also six weeks after the anterior cruciate ligament rupture operation. For each patient, the difference between the initial value immediately post-surgical and the follow-up value was determined and this difference was compared between the two study arms. This procedure has the advantage that the variance of the difference should be smaller than the variance of the IKDC score during follow-up. This makes it possible to detect smaller effects with the same number of cases. The details of the statistical analysis were defined in the statistical analysis plan (see Appendix 2 SAP CAMOped V1.1 20201214).

##### Expected effect

The expected effect of an active knee brace six weeks after surgery for a ruptured anterior cruciate ligament on the patient’s estimated knee function using the subjective IKDC was difficult to estimate based on published data. From the search in August 2020, there were no data from prospective, randomized, methodologically sound studies on rehabilitation success after surgical treatment of an ACL rupture that would have been fully suitable for calculating the number of cases. There was not a single published, methodologically superior study documenting the treatment success of CPM or CAM splints or other comparable rehabilitative measures after reconstruction of anterior cruciate ligament ruptures using the IKDC or another validated, quantifiable measurement instrument. In the study by Noll^22^ a mean IKDC value of 50.2 with a standard deviation of 9.6 was reported after four weeks for 74 patients who received physiotherapy as a rehabilitation measure following a rupture of the anterior cruciate ligament.

##### Determination of case numbers

The following hypotheses should be tested:

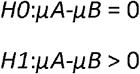

The difference between the two mean values (*μ*) in relation to the standard deviation (*σ*), the expected effect size *θ* was determined:

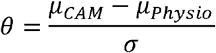

For the calculation of the number of cases and the power and significance levels, a probability of error *α* = 0,05 and a power 1 – *β* = 0,9 was assumed. The calculation was performed with R (R Core Team 2019) in version 3.6.0 and the R base package gsDesign^23^ in version 3-1-1.

The selected trial design can detect an effect of θ = 0.21 with 90% power and an α of 5%, whereby 147 patients had to be included as the expected value. With the chosen design, effects of θ>0.7 should already be detectable in the first interim analysis after 78 patients.

An effect of θ>0.7 should be detectable with 78 patients. The effect size θ can be converted into mean differences between the two treatment groups. An effect size of 0.7 with a standard deviation of 9.6 (according to Noll^22^) means a difference in the mean values of both groups of 6.7 points in the IKDC. In order to demonstrate the same effect size, there would have to be a greater difference between the mean values of the two groups with a higher standard deviation. With a standard deviation of 13.5 or 15.7 (according to Kümmel^24^), the mean values should differ by 9.5 or 11 points in the IKDC in order to be able to prove an effect size of 0.7. The assumed standard deviations covered the range of standard deviations observed in other studies. Further details on the invoice can be found in Appendix 1 Test plan RCT CAMOped V1.2 20201214, Annex 11.12.3.

The calculation was carried out with R (R Core Team 2019) in version 3.6.0 and the R-Base package gsDesign in version 3-1-1.

#### 7.2. Interim analysis

In view of the uncertainty about the expected effect, a group-sequential experimental design was appropriate. ^25^In a group-sequential experimental design, interim analyses are used to adjust the number of cases during the conduct of the study or to terminate the study if the results of the interim analysis lead to the rejection of the null hypothesis. The error probability of incorrectly rejecting the null hypothesis is kept at the predetermined level across the interim analyses and the final data evaluation without artificially increasing the error through multiple testing. Compared to trial designs with rigid case number calculation, group-sequential trial designs also have the advantage of being able to randomize an existing effect earlier in most cases. Group-sequential experimental designs have three parameters to be determined in advance:

1. the minimum number of cases,
2. the maximum number of cases,
3. the number and timing of interim analyses

A one-sided group-sequential experimental design with an initial probability of error α=0.05 and a power 1-β=0.9 was used. The minimum number of cases was set at 75 patients in order to be able to carry out an adequate descriptive statistical analysis in addition to the hypothesis test. The maximum number of cases was set at 200 patients, with interim and final analyses planned after 75, 125, 150 and 200 patients. For the interim and final analyses, adjusted critical values and significance levels were applied according to the O’Brien-Fleming function.

This approach should ensure that a larger effect of the active knee brace does not include an unnecessarily large number of patients, while smaller effects can also be detected with sufficient certainty.

### 8. Selection of the data set for the interim analysis and handling of missing values

In accordance with the planned group-sequential trial design, an interim analysis was carried out in September 2022 (see Appendix 3 Interim analysis report).

The primary analysis dataset for the interim analysis was the intention-to-treat population. This dataset contained all patients who were included in the clinical trial and randomized.

In accordance with the statistical analysis plan (see Appendix 2 SAP CAMOped V1.1 20201214), the first interim analysis was performed after the inclusion of 78 patients.

The database for conducting the interim analysis was closed after the last patient of the consecutive 78 patients had reached FU2 (6 weeks post surgery). These were randomized into the study and thus included. The database did not contain any information on the group affiliation of the study participants.

There is no database from which information on CAM treatment in combination with the sIKCD can be obtained. In order to determine a relationship between the duration of use of the CAM splint and the value of the sIKDC at T4, the unblinded team calculated a non-parametric correlation between these two variables in a separate and closed database.

Once the database for the interim analysis had been closed, the original database was stored in the data management system in an audit-proof manner (database closure).

All subsequent calculations were carried out on the basis of this audit-proof database and were versioned (including the current date) and saved in a corresponding folder in an audit-proof data management system (ecoDMS).

Missing values were not replaced in the primary analysis. The group affiliation was subsequently entered into the database in binary code by unblinded members of the MEDIACC study team, without indicating which of these numbers were assigned to the control or intervention group.

### 9. Randomisation

After recruitment and inclusion according to the inclusion and exclusion criteria, the patients were randomly assigned to the treatment arms according to the stratification characteristics (age and physical activity according to the Tegner score):

- Control group: standard physiotherapy or CPM in the subgroup with concomitant injury requiring intervention
- Intervention group: standard physiotherapy and use of CAMOped

The randomization lists according to the strata were generated by a random generator in MEDIACC for the entire study. Block randomization was performed according to the variables “Tegner score” and “age” in blocks of 10. This was to ensure that the variables considered to have the strongest influence were distributed in both treatment arms.

It was therefore impossible for unblinded MEDIACC study staff to randomize patients in any order other than that determined by the random generator.

### 10. Mechanisms for maintaining the confidentiality of treatment successes

The trial centres reported a patient to be randomized to a MEDIACC employee. Patients were then assigned to a stratum from the randomization list based on the two variables “Tegner score” and “age”. The patient’s pseudonym was entered in the randomization list in an audit-proof manner. An unblinded MEDIACC employee triggered the delivery of the investigational product (IG) if necessary or wrote to the patient to inform them of their assignment to the control group.

In accordance with the standard instructions for this study, extensive measures were taken to ensure that the group allocation was safely concealed.

This included the following measures:

- Work according to GCP and audited standard instructions of MEDIACC, defined processes after gap analysis, maintenance of a CAPA document
- Ongoing quality control and quality management

By assigning different physicians in the centres (blinded, unblinded) to specific roles, the blinding of study personnel to record endpoints during the follow-up period could be avoided even in the presence of SAEs.

In addition, patients were informed never to disclose their group affiliation and compliance with this requirement was also regularly recorded.

### 11. Blinding

Treatment providers in the study centres were always blinded and remained so for the duration of the follow-up treatments.

This was ensured by patient training, randomization by 2 trained MEDIACC employees who do not communicate the randomization result to the study centre, limited communication of patient information to MEDIACC, maintenance of the randomization lists in the study centres by trained staff. The group allocation is carried out undercover by specially designated and documented experienced, qualified and trained MEDIACC staff based on the standard instructions for this study. These employees had no role in the work, which was safely blinded. This work took place in a room with restricted access to all other MEDIACC staff and any other potential visitors in accordance with MEDIACC standard operating procedures.

In the event that a patient was assigned to the treatment group with the investigational product, the sponsor only received the details necessary for delivery.

For queries / problems of a medical nature that COULD involve the trial product, there was a trained doctor / nurse at each centre who was not involved in the rest of the patient’s treatment. Every contact was documented. In case of medical problems safely outside the use of the investigational product, patients contacted their physicians, in case of queries / problems of a technical nature they contacted the sponsor, in case of questions about the study they contacted MEDIACC (2 unblinded employees).

It was therefore impossible for employees of the manufacturer, the individual test centres or other MEDIACC employees to view the concealed group allocation.

All processes were established in MEDIACC, checked by multiple test runs and tested by trained personnel for gaps in security both internally and externally.

- Various measures were implemented and tested to ensure the blinding of the endpoint investigators:
- Quality control via scientific and regulatory knowledge of the staff in the test centres
- Work according to GCP and audited standard instructions of MEDIACC, defined processes after gap analysis, maintenance of a CAPA document
- Ongoing quality control and quality management
- Remuneration of study services via MEDIACC case flat rates, independent of success
- Exclusion of any contact of the manufacturer’s personnel with that of the test centres by the study design
- Design, testing and training of communication plans for all study staff
- Selection of trial centres with documented experience in conducting high-quality clinical trials
- Training staff in the test centres, identifying problem areas and randomized procedures
- Distribution of the endpoint survey across several information channels, different data collection tools (in particular online questionnaires for patients for all subjective parameters, demographic data, documentation of physiotherapy and training with the test product) and automated transfer of patient data to the database.
- Ensuring unblinded medical contact persons at the trial centre who have received documented training and are certainly not involved in the conduct of the trials
- Standardised prescription of a CAMOped for ALL patients in the study. The prescriptions are only sent to MEDIACC after the prescription for the last patient from each trial centre and only the prescriptions of the group that received CAMOped are forwarded to the manufacturer. The remaining prescriptions remain in MEDIACC’s TMF for documentation purposes.
- Delivery of the test products directly to the patients’ addresses
- Training of study patients by qualified personnel (manufacturer) who are not otherwise involved in the study
- Ensuring contact persons and contact details in the MEDIACC for all study matters and, if necessary, separate medical staff in the trial centres
- Personalised approach to all study patients by documented, experienced, qualified and trained MEDIACC staff who do not collect endpoints
- Safety management for AE and SAE, for which unblinded, qualified and trained personnel are available at the test centres

As can also be seen from the standard instructions for this study, extensive measures were taken to ensure that the group allocation was securely concealed.

For the interim analysis, a second randomization level was used to ensure the blinding of the evaluators. The analysis was programmed by blinded personnel and the assignment to the respective treatment groups was only carried out after all work on the audit-proof database had been completed in order to maintain the blinding of all investigators.

#### Unblinding

Unblinding of the investigators was not planned at any time. In the event of an AE or SAE caused by an investigational product, the patient was assigned a specialist contact person at the trial centre who was instructed and trained, but had no further connection to the trial.

### 12. Statistical methods

The evaluation was carried out in accordance with the explanations in the statistical analysis plan and the study protocol. The statistical methods are described below.

#### 12.1 Methods for comparing the groups (primary research question)

The intention-to-treat population was defined as the primary evaluation dataset. This data set contains all patients who were included in the clinical trial and randomized. The secondary analysis dataset is the per-protocol population. This dataset contains all patients who were treated according to the protocol for the entire duration of the study. The tertiary analysis dataset (safety population) contains all patients who received the study treatment. The composition of the study population was described in detail in Chapter 4.2.1 of the protocol (see Appendix 1 RCT protocol CAMOped V1.2 20201214).

##### The randomizations for the per protocol analysis and the tertiary data set are not shown below

All analyses here were carried out using data from all patients who could be assigned to the intention-to-treat population. According to the study protocol, treatment within the study was considered intended as soon as a patient was randomized.

In this analysis, all available data are presented without taking into account missing data. The patients contributed with the available data up to the time of leaving or completing the study.

The primary endpoint of change in joint function measured by IKDC was analysed after 6 weeks of treatment with CAMOped compared to standard therapy.

###### Primary analysis

The primary endpoint is the change in knee joint function as measured by the subjective IKDC. It was analysed 6 weeks post surgery (FU2) in relation to the immediate post-surgical assessment (T4) under therapy with CAMOped compared to standard therapy.

The subjective knee joint function was calculated according to the “Instructions for calculating the result for the 2000 form for the subjective assessment of the knee” contained in the IKDC. The individual items of the sIKDC were scored according to the instructions and the total was calculated. A maximum score of 87 points can be achieved. This score was then transformed onto a scale from 0 to 100 in accordance with the instructions.

#### 12.2 Secondary analyses, subgroups, further analyses

The subgroup of patients with concomitant knee injuries requiring intervention diagnosed intraoperatively should be analysed separately per protocol.

##### Predefined subgroup analysis of ACLR with concomitant injuries

Subgroup of patients with concomitant knee injuries requiring intervention diagnosed intraoperatively:

Primarily patients with isolated ACLR should be included in the CAMOPED study, as more extensive operations have a significant impact on the subsequent rehabilitation programme and thus on the primary endpoint after 6 weeks. ^26^However, despite careful planning of the operation, more extensive knee findings requiring intervention may be present intraoperatively (see also Appendix 1 RCT CAMOped V1.2 trial protocol 20201214, Section 1.5.6.2), which must be treated unilaterally in accordance with the guidelines. In order to keep the present study with ITT approach meaningful, patients should continue to be followed up in the study in the event of a more extensive intraoperative treatment, e.g. a meniscus, and still receive the guideline-compliant treatment. At some centres, this included treatment with CPM splints as standard in addition to physiotherapy and it was considered unethical to withhold CPM splints from these patients – despite the fact that the G-BA had not yet made a decision at the time.

These patients were to be analysed separately in a per-protocol analysis as a subgroup in addition to the patients in the main arm of the study.

There are several reasons for the completely separate evaluation of patients who receive an additional e.g. meniscus operation:

- All patients who require further intraoperative interventions with a divergent rehabilitation programme, e.g. after meniscus reconstruction, form a different disease entity that is not comparable with the other patients.
- Nevertheless, due to the frequency of occurrence, patients with concomitant meniscal injuries are also part of the target population. It is therefore possible to extend the indication for CAMOped from isolated ACLRs to ACLRs with concomitant injuries.
- ACLR patients with concomitant injuries receive a randomize mobility splint (CPM) as standard in addition to physiotherapy; this is considered a distorting mandatory exclusion criterion in the main group. The CAM splint can therefore not be compared with physiotherapy alone.
- To date, there are no results from a study of comparable methodological quality to this one for patients undergoing ACLR surgery with concomitant injuries for the use of CPM splints worldwide. Furthermore, there are no results worldwide on the effectiveness of CPM splints after ACLR in which a pre-specified endpoint was recorded using a validated survey instrument (IKDC). A comparison of the efficacy of CAMOpeds with that of CMP splints can therefore only be made in a study in which CPM splints are also assessed using the same survey instruments. It was therefore possible to investigate whether CAMOped is at least non-inferior to CPM splints.

For patients from the subgroup with ACLR with concomitant injuries, 2 comparison arms were defined according to the previously performed and remaining randomization:

A. Patients who have received a CAMOped as a result of preoperative randomization continue to use it.
B. Patients from the original control group receive a CPM splint.

Patients who intraoperatively fall into the subgroup of patients with concomitant injuries should be replaced in the main arm by newly recruited patients until the number of cases for the main arm could be reached.

#### 12.3 General procedure

Descriptive statistics were used to summarise the endpoints and baseline characteristics.

Descriptive statistics such as mean, median, standard deviation and quartiles were used to summarise continuous variables. Dichotomous variables were presented in numbers and percentages.

The health-related quality of life was calculated on the basis of the points indicated on the Likert scale, from which the total score was calculated.

All continuous variables were analysed for normal distribution using a Q-Q plot. Normally distributed data were represented as mean ± standard deviation, non-normally distributed by median and interquartile range.

The Mann-Whitney-U test was used to test the superiority of the CAM splint over physiotherapy.

Linear regression analysis was used to analyse which variables were independent predictors of subjective knee assessment post surgery (T4). For this purpose, the variables age, gender, Tegner score and graft type were included in the model.

Statistical significance was assumed if the null hypothesis could be rejected with a significance level of p ≤ 0.05. SPSS® (version 29) was used for the analysis.

Statistical principles for clinical studies according to ICH-GCP were followed.

## III Results

### 13. Number of study participants

The following Illustration *4* shows the flow chart of the clinical study.

**Illustration 4.**
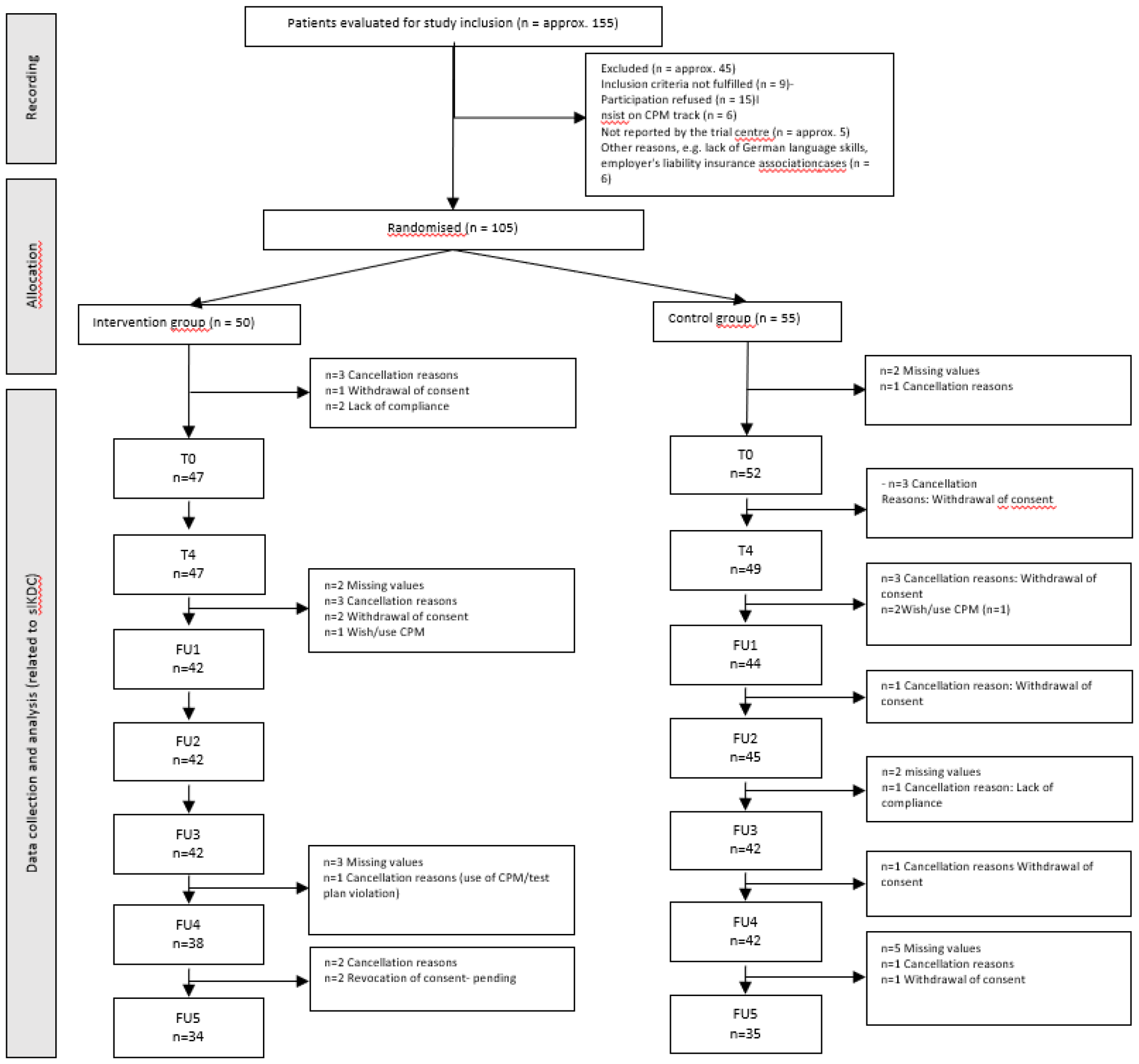
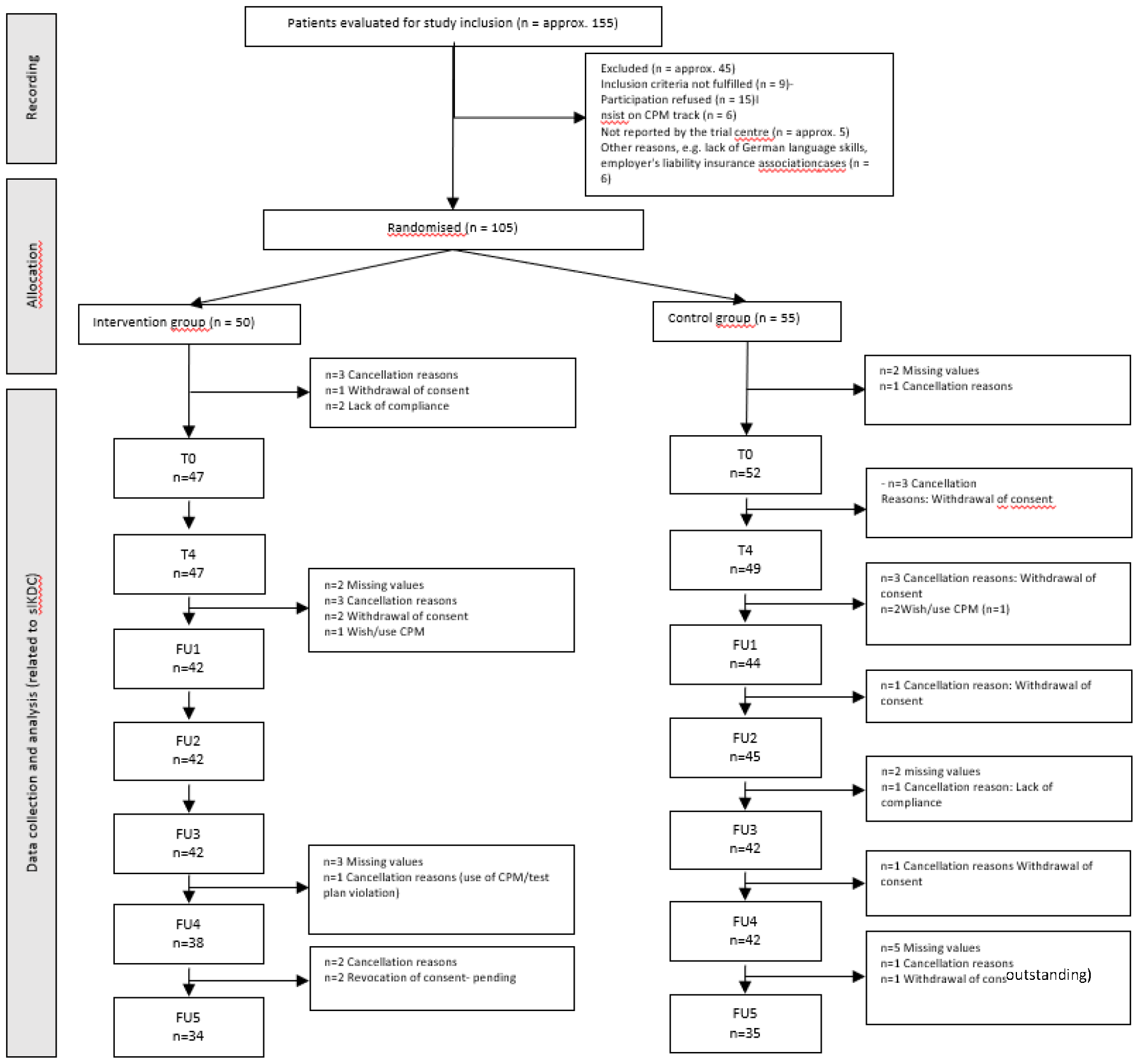
Study flow chart. Abbreviations: FU: Follow-Up; n: Number.

Six people dropped out of the study before completing the inclusion patient questionnaire for the subjective assessment of knee function (IKDC 2000).

### 14. Admission / Recruitment

#### 14.1 Time points for inclusion and follow-up observations

Between June 2020 and January 2023, around 155 patients were approached and 105 were included in the study after checking the inclusion and exclusion criteria and obtaining consent, and were treated with CAMOped in addition to standard therapy or standard therapy alone. The follow-up examinations took place at 3, 6, 12, 26 and 52 weeks post surgery.

#### 14.2 Date and reason for the end of the study programme

From 12/2021 (after the CPM splint was included in standard care, BAnz AT 04.09.2019 B2), the high number of refusals to participate in a study without CPM splints in the control group by patients and the number of protocol violations and dropouts were striking.

The recruitment performance of the study had been constant, interrupted by the legal measures during the corona pandemic with effects on the possibilities to perform injury-prone sports and the restrictions in the respective clinics at a low level, as shown in Illustration 5A is shown. In Illustration 5B shows the reasons for exclusion.

**Illustration 5.**
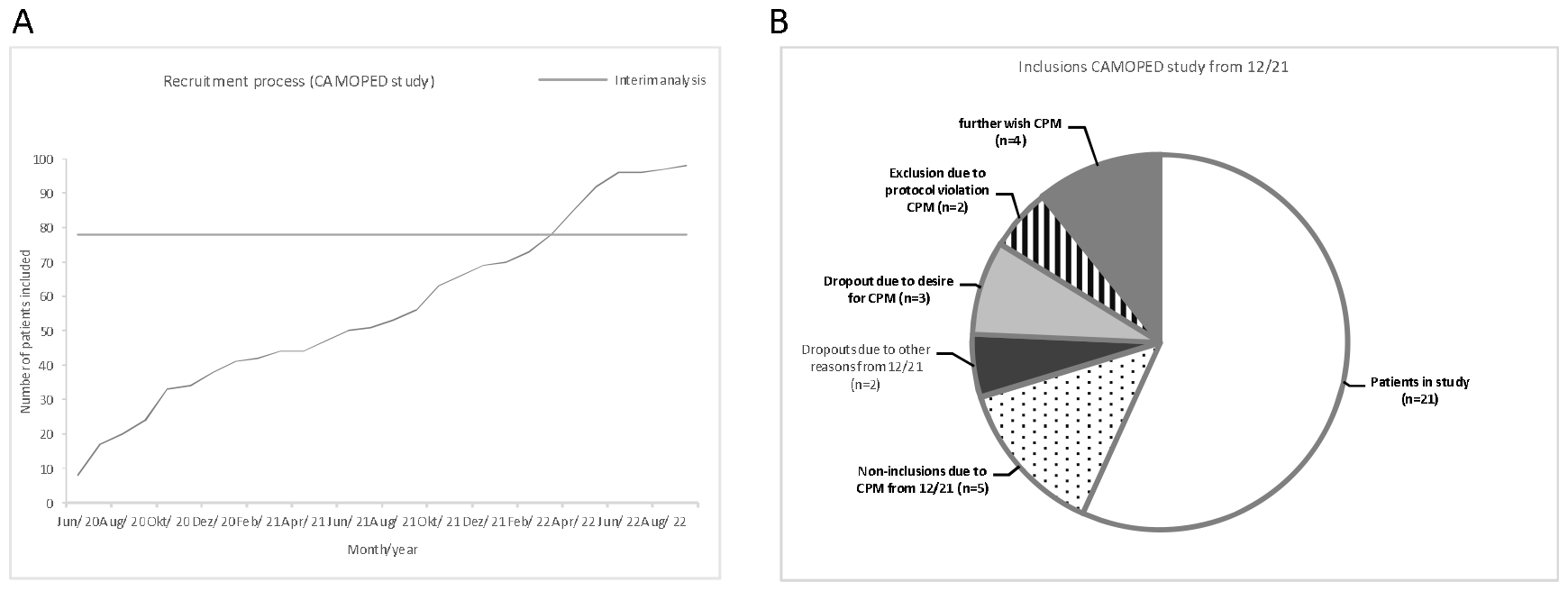
A Recruitment process during the study, B Reasons for exclusion from 12/2021 to the time of the interim analysis.

As shown in Illustration 5B, only 25 of the 32 patients who have been included since December 2021 have remained in the study. This corresponds to an unusually high dropout rate of 28% over an average of only 4 months. Prior to this period (12/2021), there were 9 dropouts out of 69 patients included (13%) over 12 months each. The reasons for dropout of the 7 of the 32 patients after 12/2021 were predominantly the desire or the already existing use of a CPM splint (exclusion criterion, except for the control group with additional meniscus interventions). In addition, 5 patients in one centre refused to participate in the study due to the risk of not receiving a CPM splint. Furthermore, when asked, we received information that approx. 30% of the patients approached refused to participate in the study, in each case for the reason that they feared that they would not receive an active or passive motion splint in the event of randomisation to the CG.

Of the recruited patients from the period, 4 patients expressed the wish to use a CPM splint in addition, if necessary, or had already used a CPM splint briefly but could be kept in the study. Otherwise, there were only 2 patients in the period who discontinued the studies (longer journey and non-compliance) for other reasons.

After conducting the interim analysis on 19 September 2022 (see Appendix 3 Interim analysis report) and subsequent discussion with the LKP and the principal investigators, this led to the principal investigator terminating the study for ethical reasons (see letter from the LKP dated 23 January 2023, Appendix 4).

### 15. Demographic data

Of the 105 patients included in the study, 50 were recruited to the intervention group (IG) and 55 to the control group (CG) (Table 4). A total of 17 of the 105 patients were in the subgroup of ACLR with additional injury to the meniscus requiring intervention.

**Table 4.**
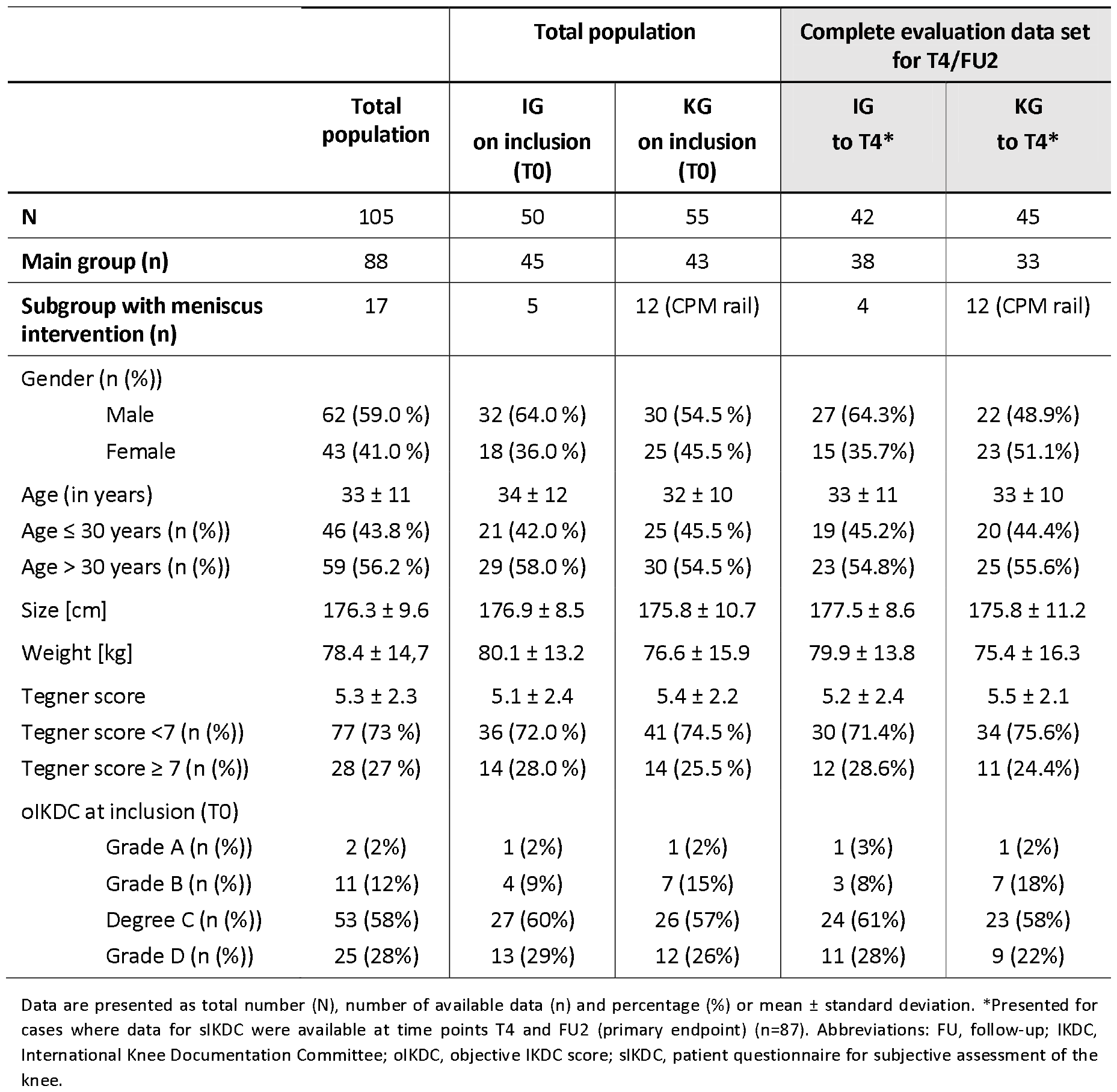
Demographic data of the study population at inclusion (T0)

In the overall study population, more men than women were included (59%: 41%). The average age was approx. 33 ± 11 years. The activity index (Tegner score) at randomisation was 5.3 ± 2.3 in the overall study population and was identical in the two treatment groups. In accordance with the stratification strategy, people aged ≤ 30 years and over 30 years and with a Tegner score < 7 or ≥ 7 were evenly distributed between the treatment groups.

Preoperatively (T0), 58% of patients in the overall population had oIKDC grade C. The proportionate distribution of oIKDC grades was comparable between patients in the IG and KG at inclusion; they had almost equal proportions of oIKDC grade C in both the overall study population and the evaluation dataset (overall study population: IG: 60%, KG: 57%; evaluation dataset: IG: 61%, KG: 58%).

Randomisation with equal distribution of the confounding variables was therefore successful. In addition, the study collective is a representation of the target population.

### 16. Number of patients analysed

At T0, information from the IKDC 2000 patient questionnaire (sIKDC) was available for 99 people. Three more people dropped out of the study between inclusion and surgery (N=96 at time T4). At time FU1, data was available from 86 patients, at time FU2 from 87 patients, 12 weeks post surgery (FU3) from 84 patients, after 6 months (FU4) from 80 patients and one year post surgery (FU5) from 69 patients. For the evaluation of the primary endpoint, 87 complete analysable data sets were available at the time points T4 and FU1 (evaluation data set).

Out of 99 patients, a total of 12 people in the control group received a CPM splint as per protocol after additional surgical intervention of the meniscus. This left only 45 patients without additional meniscus intervention in the IG and 43 in the KG for the evaluation without receiving a CPM splint (see Table 4).

### 17. Evaluation of the primary endpoint

#### 17.1 Presentation of subjective knee joint function in the course of rehabilitation for the overall population

The subjective assessment of knee joint function was recorded using the subjective patient questionnaire IKDC 2000 at the time of inclusion (T0) and at the various examination times. In each case, the sIKDC score was calculated and scaled to a range between 0 and 100, with a higher score reflecting better subjectively assessed knee function.

post surgery (time point T4), the sIKDC score in the overall population decreased as expected compared to the baseline values recorded at the inclusion time point T0 (T0: 52.0 [38.0 - 63.0] vs. T4: 30.0 [22.5 - 38.5], Table 5 and Illustration 6 A). In the course of rehabilitation, the sIKDC score increased again and was slightly higher than the initial value 6 weeks post surgery (55.0 [48.0 - 64.0]). Twelve weeks post surgery (FU3), the median sIKDC score of 69.0 [62.0 - 76.0] was higher than before the operation. In the further course of rehabilitation, the sIKDC score continued to rise and reached a median value of 84.0 [74.0 - 92.0] in the overall population one year post surgery.

**Table 5.** Results of the subjective IKDC over the study period up to FU5 for the total population and breakdown into IG and KG (all cases)

| Timing | Total population |  | IG |  | KG |  | Significance level<br>Group difference |
| --- | --- | --- | --- | --- | --- | --- | --- |
|  | N | Median [IQR] | N | Median [IQR] | N | Median [IQR] |  |
| **T0** (inclusion) | 99 | 52.0 [38.0 - 63.0] | 47 | 53.0 [40.0 - 63.0] | 52 | 51.5 [37.0 - 62.0] | n.s. |
| **T4** (1-2 days post surgery) | 96 | 30.0 [22.5 - 38.5] | 47 | 32.0 [23.0 - 46.0] | 49 | 29.0 [22.0 - 34.0] | n.s. |
| **FU1** (3 weeks post surgery) | 86 | 44.5 [37.0 - 51.0] | 42 | 46.0 [40.0 - 51.0] | 44 | 42.0 [33.5 - 48.5] | n.s. |
| **FU2** (6 weeks post surgery) | 87 | 55.0 [48.0 - 64.0] | 42 | 56.5 [48.0 - 64.0] | 45 | 54.0 [48.0 - 63.0] | n.s. |
| **FU3** (12 weeks post surgery) | 84 | 69.0 [62.0 – 76.0] | 42 | 68.0 [60.0 - 74.0] | 42 | 69.5 [63.0 - 77.0] | n.s. |
| **FU4** (6 months post surgery) | 80 | 77.0 [68.5 - 84.0] | 38 | 77.5 [69.0- 84.0] | 42 | 76.5 [68.0 - 84.0] | n.s. |
| **FU5** (12 months post surgery) | 69 | 84.0 [74.0 - 92.0] | 34 | 80.5 [70.0 - 92.0] | 35 | 85.0 [75.0 – 92.0] | n.s. |

Data are presented as median [IQR 25-75]; abbreviations: FU, follow-up; IQR, inter-quartile range; N, number; T, time; sIKDC, subjective patient questionnaire for the assessment of the knee.

**Illustration 6.**
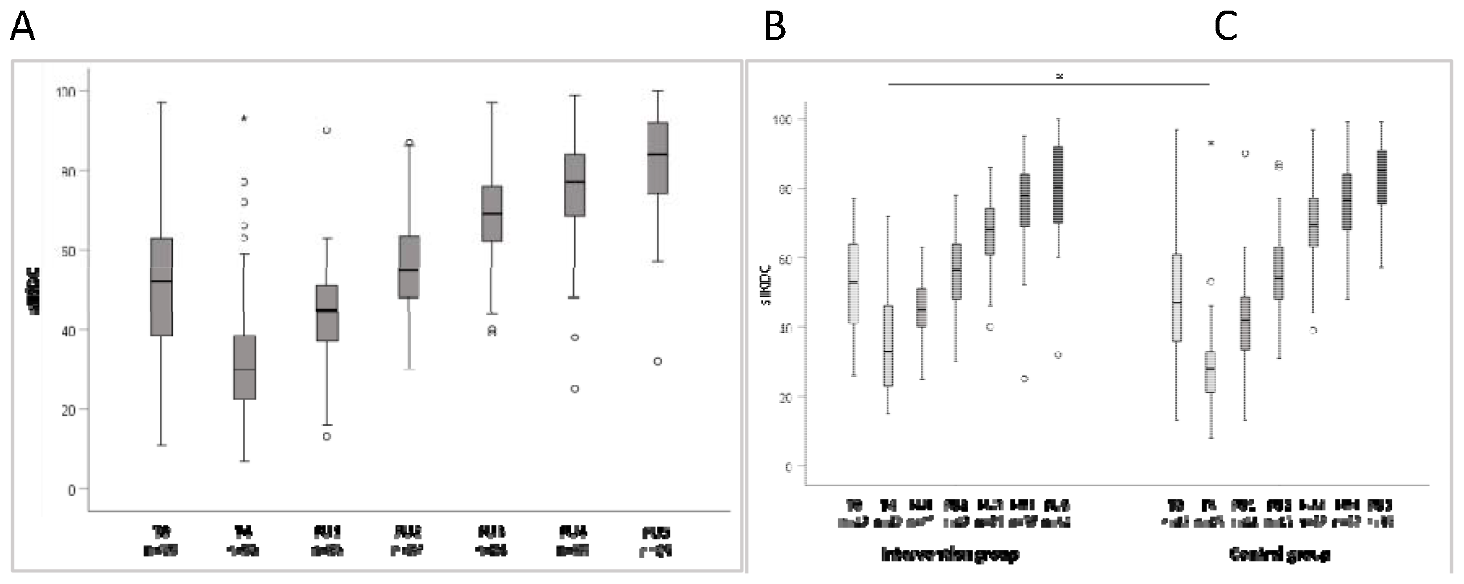
Subjectively assessed knee joint function (sIKDC score) at the various examination times in the A total population (total data set), B intervention group and C control group (evaluation data set). T0, inclusion; T4, 1-2 days post surgery ; FU1, 3 weeks post surgery ; FU2, 6 weeks post surgery ; FU3, 12 weeks post surgery ; FU4, 6 months post surgery ; FU5, 1 year post surgery ; (A: total data set; B, C: cases with available data at time points T4 and FU2). Abbreviations: FU, follow-up, T, time point; sIKDC, subjective assessment of knee joint function, * p<0.05 in group comparison.

Thus, the course of the assessment of knee joint function corresponds to the expected course of the target population.

The course of the sIKDC score was comparable in the treatment groups. Including all values of the total population (total data set), the sIKDC score was not significantly different between the treatment groups at any time (Table 5)).

At the time points of the follow-up examinations (FU1 to FU5), the values of the sIKDC score were similar between the IG and KG. The values are shown for all patients, including those with meniscus interventions and thus those in the control group (n=12) who received a CPM splint.

In the subjective assessment of post-surgical knee joint function between IG and KG at time T4 - before the start of treatment with the test product - lower values were found in the KG (Table 5), which had a higher number of patients with simultaneous meniscus intervention (evaluation data set for the primary endpoint, IG:KG 5:12, corresponding to 10% and 22% per group) with otherwise identical patient-related variables (see Table 1). As expected, patients with additional meniscal intervention had a lower sIKDC post-surgical compared to isolated ACLR (p=0.05, Table 6).

**Table 6.** Results of the subjective IKDC at time point T4 (1 to 2 days post surgery) in the skin group (isolated ACLR) and secondary group (ACLR with concomitant injuries) (evaluation data set with complete data for both time points - time points T4 and FU2, n=87)

| Timing | Isolated ACLR |  |  |  |  |  | ACLR + meniscus intervention |  |  |  |  |  |
| --- | --- | --- | --- | --- | --- | --- | --- | --- | --- | --- | --- | --- |
|  | Total |  | IG |  | KG |  | Total |  | IG |  | KG |  |
|  | N | Median [IQR] | N | Median [IQR] | N | Median [IQR] | N | Median [IQR] | N | Median [IQR] | N | Median [IQR] |
| **T4** (1-2 days post surgery) | 71 | 32.0 [23.0 - 38.0]* | 38 | 33.0 [23.0 - 47.0] | 33 | 28.0 [24.0 - 33.0] | 16 | 27.0 [19.0-30.0]* | 4 | 29.5 [25.0-35.5] | 12 | 23.5 [16.5-30.0] |

Data are presented as median [IQR 25-75]; abbreviations: IQR, inter-quartile range; N, number; T, time point; sIKDC, subjective patient questionnaire for the assessment of the knee; IG, intervention group; KG, control group. \* $p=0,05$

In a multivariate model (regression, listwise exclusion), none of the other possible influencing variables on the subjective post-surgical knee assessment (T4) were identified as independent variables. Age, gender, Tegner score and graft type were included in the model.

##### Summarised assessment (total population)

As can be seen from the equal distribution in the preoperatively relevant variables influencing the endpoint between the two treatment groups at inclusion (T0), randomisation was successful in relation to the overall population. The higher sIKCD post surgery for the subjective assessment of knee joint function in the IG compared to the KG can be explained by the approximately twice as high relative proportion of additional meniscus operations with consecutively impaired post-surgical knee joint function in the KG. In the further follow-up period, the subjective assessments of knee joint function were identical in the groups. The target population was mapped in this study.

#### 17.2 Primary endpoint: change in subjectively assessed knee joint function immediately post-surgical up to 6 weeks rehabilitation - evaluation for complete data sets at T4 and FU2

The primary objective of the study was to examine whether rehabilitation with CAMOped in addition to standardised rehabilitation is superior to standardised rehabilitation alone after 6 weeks post-surgical following surgical reconstruction of a unilateral rupture of the anterior cruciate ligament (ACL), in terms of knee joint function as measured by sIKDC. All values of the total population for which data on sIKDC at T4 and FU2 were available were used for this analysis. From 87 (IG: 42, KG: 45) patients, values were available both at T4 (post surgery) and 6 weeks after surgery (FU2). There were 38 patients in the IG without additional meniscus intervention and 33 in the KG. In the CG, a total of 12 people received a CPM splint with additional surgical intervention of the meniscus. The data for the 87 patients is presented below, even though 12 of them used a CPM splint in accordance with the protocol and were not a control group in the original sense without the use of a motion splint.

When analysing the cases for which sIKDC values were available both at T4 and at FU2 (time of the primary endpoint), the sIKDC score in the IG at inclusion (T0) was comparable to that in the KG (Table 7, Illustration 6 B,C). The sIKDC score post surgery (T4) in this evaluation set was significantly higher in the IG than in the CG (33 [23.0 - 46.0] vs. 28 [21.0 - 33.0], p=0.024, for explanation see Table 6).

**Table 7.**
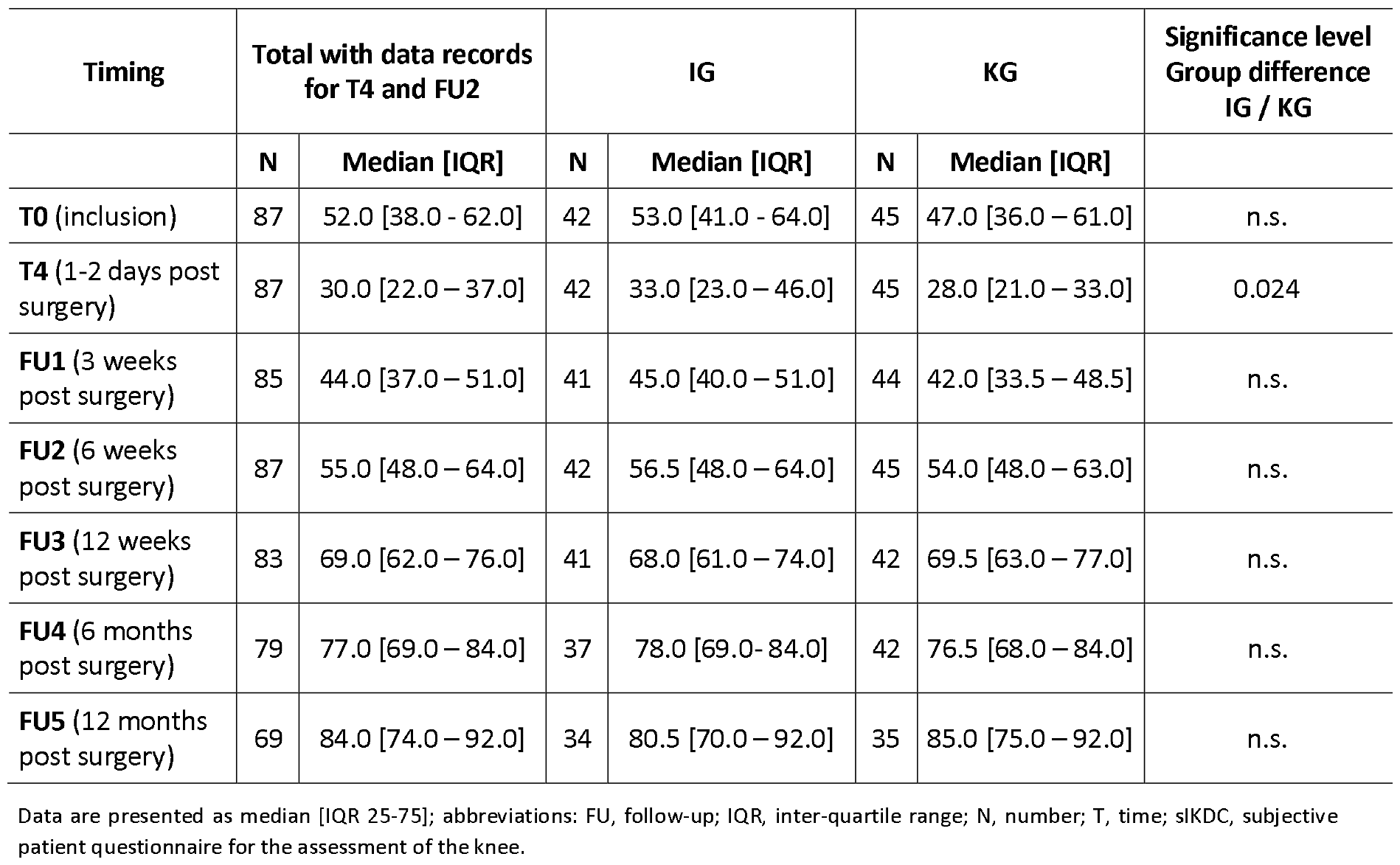
Results of the subjective IKDC over the study period up to FU5 for the total population and the treatment groups (evaluation data set with complete data sets for time points T4 and FU2)

| Timing | Total with data records for T4 and FU2 |  | IG |  | KG |  | Significance level Group difference IG / KG |
| --- | --- | --- | --- | --- | --- | --- | --- |
|  | N | Median [IQR] | N | Median [IQR] | N | Median [IQR] |  |
| **T0** (inclusion) | 87 | 52.0 [38.0 - 62.0] | 42 | 53.0 [41.0 - 64.0] | 45 | 47.0 [36.0 - 61.0] | n.s. |
| **T4** (1-2 days post surgery) | 87 | 30.0 [22.0 - 37.0] | 42 | 33.0 [23.0 - 46.0] | 45 | 28.0 [21.0 - 33.0] | 0.024 |
| **FU1** (3 weeks post surgery) | 85 | 44.0 [37.0 - 51.0] | 41 | 45.0 [40.0 - 51.0] | 44 | 42.0 [33.5 - 48.5] | n.s. |
| **FU2** (6 weeks post surgery) | 87 | 55.0 [48.0 - 64.0] | 42 | 56.5 [48.0 - 64.0] | 45 | 54.0 [48.0 - 63.0] | n.s. |
| **FU3** (12 weeks post surgery) | 83 | 69.0 [62.0 - 76.0] | 41 | 68.0 [61.0 - 74.0] | 42 | 69.5 [63.0 - 77.0] | n.s. |
| **FU4** (6 months post surgery) | 79 | 77.0 [69.0 - 84.0] | 37 | 78.0 [69.0 - 84.0] | 42 | 76.5 [68.0 - 84.0] | n.s. |
| **FU5** (12 months post surgery) | 69 | 84.0 [74.0 - 92.0] | 34 | 80.5 [70.0 - 92.0] | 35 | 85.0 [75.0 - 92.0] | n.s. |

Data are presented as median [IQR 25-75]; abbreviations: FU, follow-up; IQR, inter-quartile range; N, number; T, time; sIKDC, subjective patient questionnaire for the assessment of the knee.

The absolute difference in the sIKDC score between 1-2 days post surgery (T4) and 6 weeks post surgery was 29.0 [15.0 - 32.0] in the data set of patients in this evaluation (Table 8). The absolute difference of the sIKDC score in the IG was significantly lower than that of the sIKDC score in the CG (24.5 [12.0 - 31.0] vs. 29.0 [18.0 - 35.0], p=0.041). Accordingly, the relative change in subjective knee joint function 6 weeks post surgery (FU2) in relation to the initial findings 1-2 days post surgery was also significantly lower in the IG compared to the CG (0.42 [0.19 - 0.57] vs. 0.53 [0.37 - 0.60], p=0.039) (Table 8). Reasons for the lower value of sIKDC at T4 in the CG can be found in chapter 17.1 and Table 6.

**Table 8.** Absolute and relative change in subjectively assessed knee joint function (sIKDC) from 1 to 2 days post surgery (T4) to 6 weeks post surgery (FU2) in IG and KG.

| Timing | Total population |  | IG |  | KG |  | p-value<br>[IG vs. KG] |
| --- | --- | --- | --- | --- | --- | --- | --- |
|  | N | Median [IQR] | N | Median [IQR] | N | Median [IQR] |  |
| **T4 to FU2**<br>Absolute difference | 87 | **29.0** [15.0 – 32.0] | 42 | **24.5** [12.0 - 31.0] | 45 | **29.0** [18.0 – 35.0] | 0.041 |
| **T4 to FU2**<br>Relative change | 87 | **49%** [29% - 58%] | 42 | **42%** [19% - 57%] | 45 | **53%** [37% - 60%] | 0.039 |

Abbreviations: FU, Follow-Up; T, Time.

##### Summarising the evaluation of the primary endpoint

A significant difference in the change in subjective knee joint function 6 weeks post surgery compared to baseline was shown in both absolute and relative (primary variable) values between the groups.

The subjective assessment of knee joint function did not differ significantly at inclusion, but did differ significantly at baseline (T4, a few days post surgery) between the (IG: 33 [23.0 - 46.0] vs. KG: 28 [21.0 - 33.0], p=0.024), cf. Table 7. Six weeks (FU2) post surgery, there was no statistically significant difference in the sIKDC score between the treatment groups. The difference in the absolute and relative change in the subjectively assessed knee joint function after 6 weeks (FU2) in relation to the post surgery (T4) value was therefore largely due to the significantly higher value at T4. This was most likely a consequence of the only half as high proportion of additional interventions for meniscus injuries in this group. As the surgeons could not have known about the group allocation, this must be an unequal distribution with too small a number of cases.

#### 17.3 Influence of CAMOped use on knee-related quality of life

Knee-related quality of life was assessed using the subscale of the patient questionnaire KOOS (Knee Injury and Osteoarthritis Outcome Score; questions Q1 to Q4) at the time of enrolment (T0) and at the various examination times. The KOOS score (QoL) was calculated in each case and scaled to a range between 0 and 100, with a higher score reflecting a better knee-related quality of life.

Post-surgical, the KOOS (QoL) score in the overall population decreased from the baseline value at inclusion of 25.0 [12.5 - 37.5] to 12.5 [0.0 - 25.0] (Illustration 7). In the course of rehabilitation, knee-related quality of life increased continuously and was 31.3 [12.8 - 43.8] higher than the baseline value 6 weeks post surgery (FU2). One year after the operation, the KOOS QoL score reached a median value of 68.8 [50.0 - 81.3] and thus corresponds to the values of the target population.

**Illustration 7.**
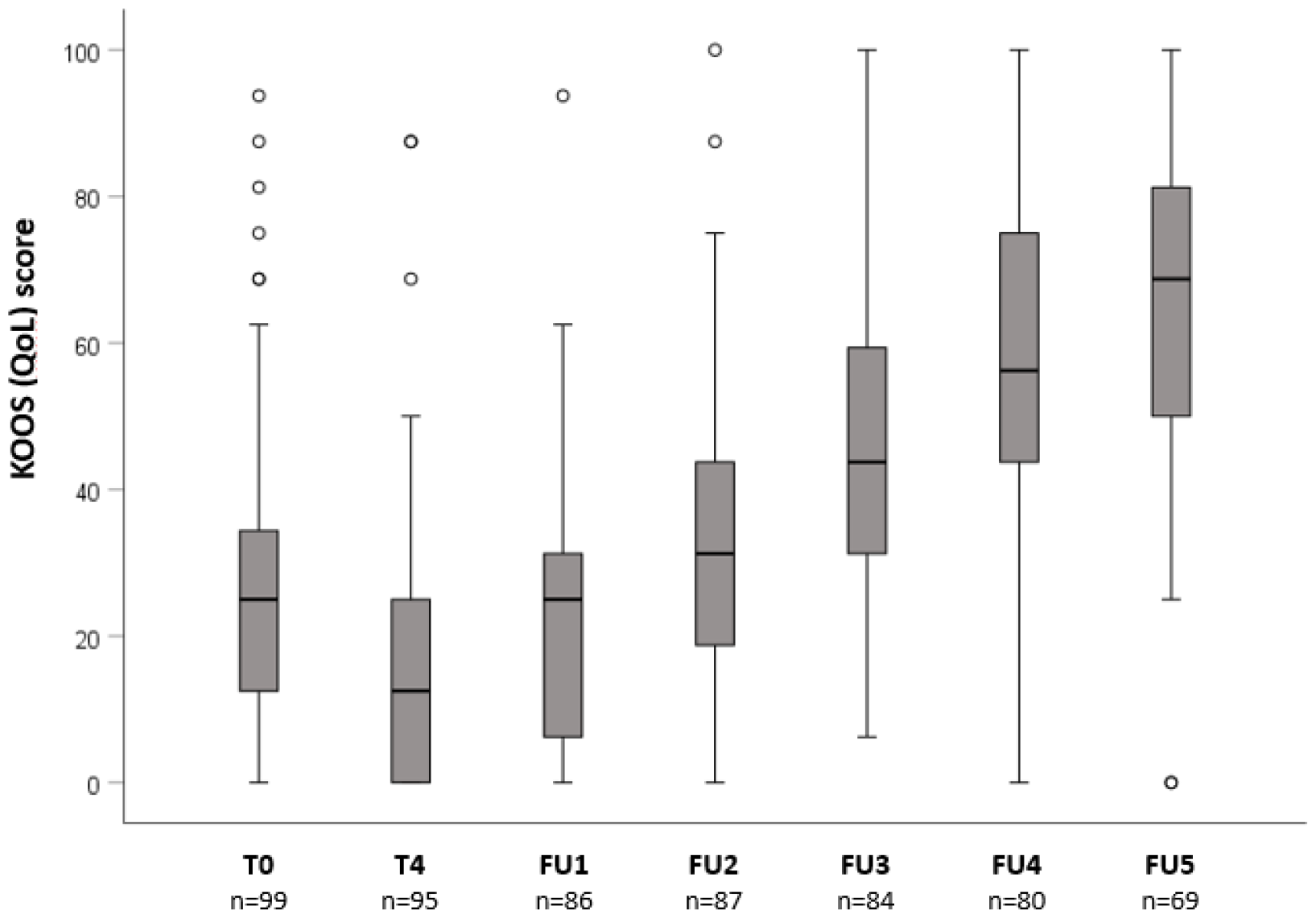
KOOS (QoL) score at the different study time points in the total population. Abbreviations: FU, Follow-Up; KOOS, Knee Injury and Osteoarthritis Outcome Score; QoL, Quality of Life; T, Time.

When analysing the cases for which values were available both at time point T4 and at FU2 (time point of the primary endpoint, analysis dataset), the KOOS QoL score was not significantly different between the treatment groups at any time point. In particular, there were no differences in the KOOS QoL score between the treatment groups at T4 (Illustration 8, Table 9).

**Table 9.** Results of the KOOS (QoL) score over the study period up to FU5 for the total population and the treatment groups (evaluation group with complete data sets for time points T4 and FU2)

| Timing | Total population |  | IG |  | KG |  | Significance level<br>Group difference |
| --- | --- | --- | --- | --- | --- | --- | --- |
|  | N | Median [IQR] | N | Median [IQR] | N | Median [IQR] |  |
| **T0** (inclusion) | 86 | 25.0 [12.5 – 31.3] | 41 | 25.0 [12.5 – 31.3] | 45 | 18.8 [12.5 – 31.3] | n.s. |
| **T4** (1-2 days post-surgery) | 86 | 12.5 [0.0 – 25.0] | 41 | 12.5 [0.0 – 25.0] | 45 | 12.5 [6.3 – 25.0] | n.s. |
| **FU1** (3 weeks post-surgery) | 84 | 25.0 [9.4 – 31.3] | 40 | 25.0 [9.4 – 31.3] | 44 | 25.0 [9.4 – 31.3] | n.s. |
| **FU2** (6 weeks post-surgery) | 86 | 31.3 [18.8-43.8] | 41 | 31.3 [18.8-43.8] | 45 | 31.3 [25.0 – 50.0] | n.s. |
| **FU3** (12 weeks post-surgery) | 82 | 43.8 [31.3 – 62.5] | 40 | 43.8 [31.3 – 56.3] | 42 | 43.8 [31.3 – 62.5] | n.s. |
| **FU4** (6 months post-surgery) | 78 | 56.3 [43.8 – 75.0] | 36 | 62.5 [43.8 – 75.0] | 42 | 56.3 [43.8 – 75.0] | n.s. |
| **FU5 (12 months post-surgery)** | 69 | 68.8 [50.0 – 81.3] | 34 | 65.6 [50.0 – 75.0] | 35 | 68.8 [56.3 – 87.5] | n.s. |

Abbreviations: FU, follow-up; IQR, inter-quartile range; N, number; T, time; KOOS, Knee Injury and Osteoarthritis Outcome Score; QoL, Quality of Life.

**Table 10.**
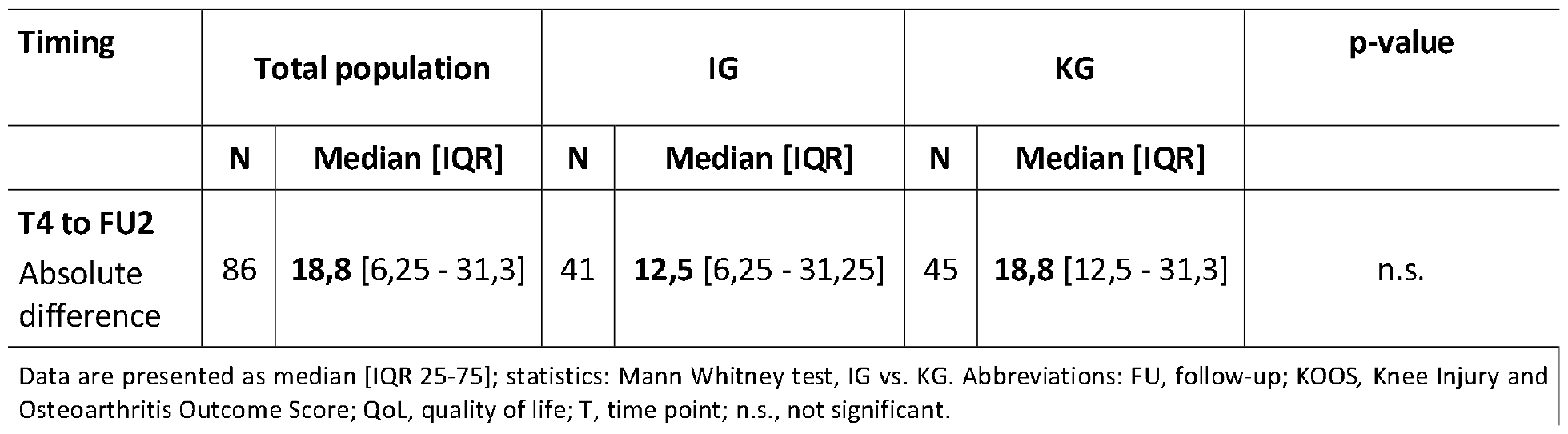
Change in KOOS from 1 to 2 days post surgery (T4) to 6 weeks post surgery (FU2) in the total population.

**Illustration 8.**
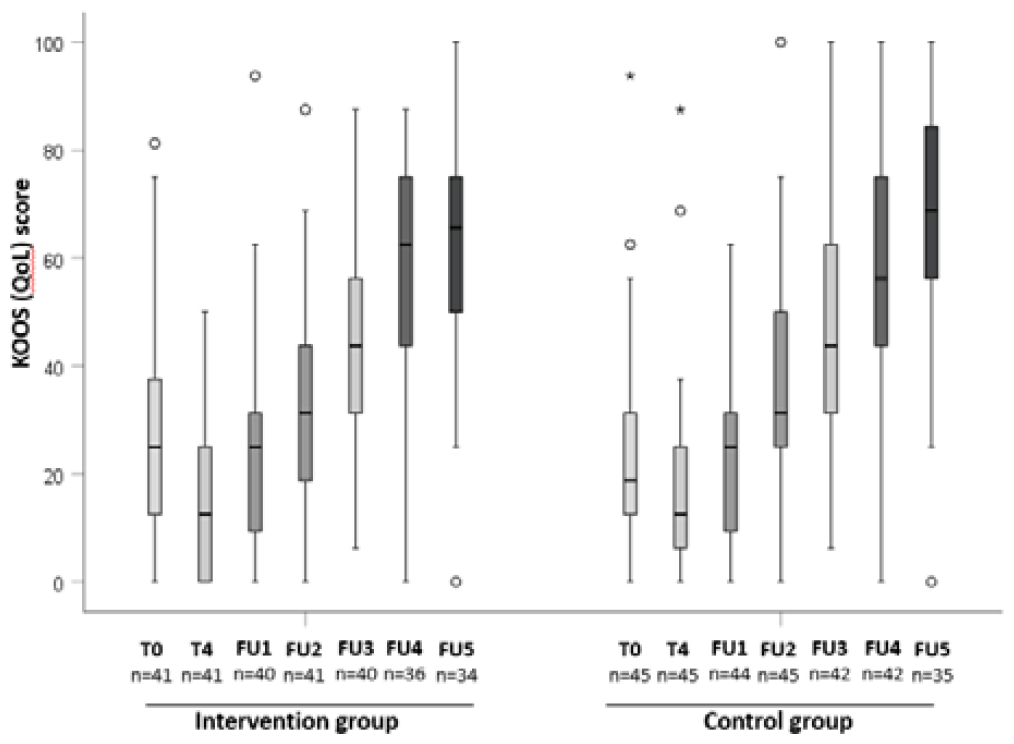
Knee-related quality of life (KOOS QoL score) at the various examination times in the respective examination groups. T0, inclusion; T4, 1-2 days post surgery ; FU1, 3 weeks post surgery ; FU2, 6 weeks post surgery ; FU3, 12 weeks post surgery ; FU4, 6 months post surgery ; FU5, 1 year post surgery); analysis dataset (cases with available data at time points T4 and FU2); abbreviations; FU, follow-up; n, number; T, time point; KOOS, Knee Injury and Osteoarthritis Outcome Score; QoL, Quality of Life.

The further calculations provided for in the statistical analysis plan (percentage change in the sum of the sIKDC score and the KOOS-QoL score from T4 to time FU2) were not carried out due to the premature termination of the study and the resulting insufficient significance of the study.

### 18. Additional analyses

#### 18.1 Progression of the objective IKDC

The objective IKDC corresponded to the expected values of the target population and, as described in the preparation of the protocol, is not suitable for describing the course of rehabilitation in relation to a treatment option. Preoperatively, the majority of patients had grade C. Post surgery, all patients had grade D. Over the course of the programme, the assessment of the oIKDC improved, as shown in Illustration 9 shown in Figure 9. There was no difference between IG and KG and no significant difference between isolated ACLR intervention and ACLR with meniscus intervention.

**Illustration 9.**
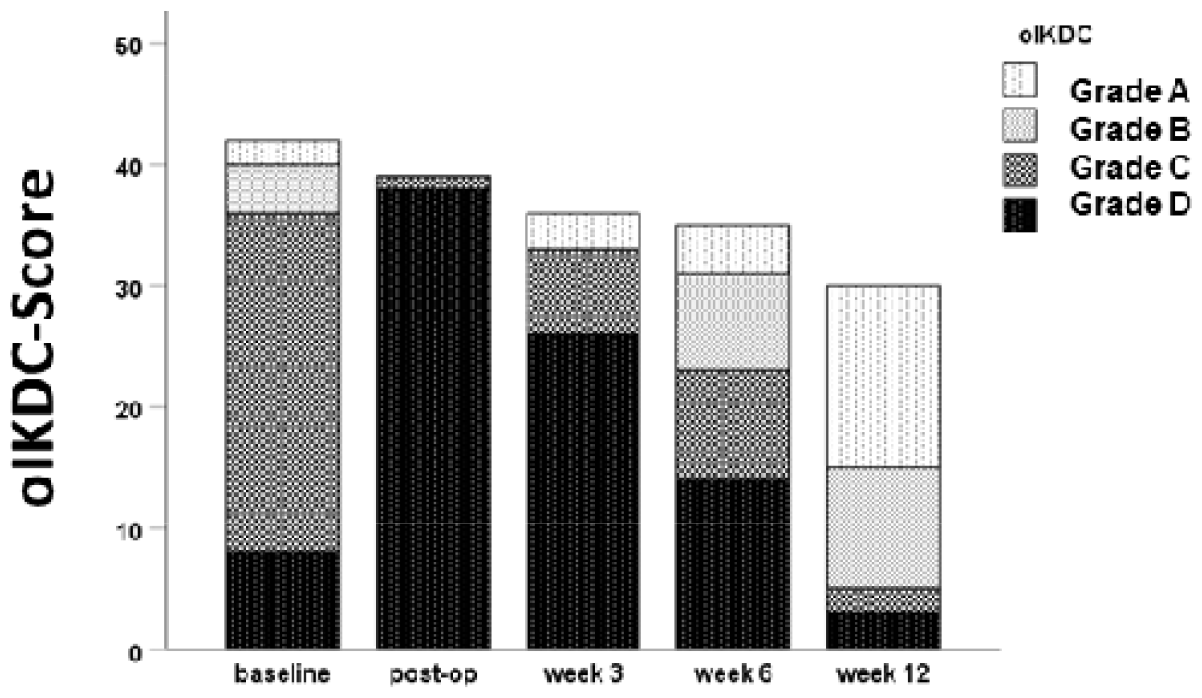
Objective IKDC score [0-87 points] over 12 weeks Classification grade A-D corresponds to: A = normal, B = almost normal, C = abnormal, D = clearly abnormal

#### 18.2 Use of CAMOped and utilisation of various rehabilitation measures

Due to the premature termination of the study, a final evaluation of these parameters with all included patients was omitted. The results of an interim analysis are shown in Illustration 10 and Illustration 11 are shown. Illustrations and descriptions are taken from the dissertation by W. Karaszewski.^27^

**Illustration 10.**
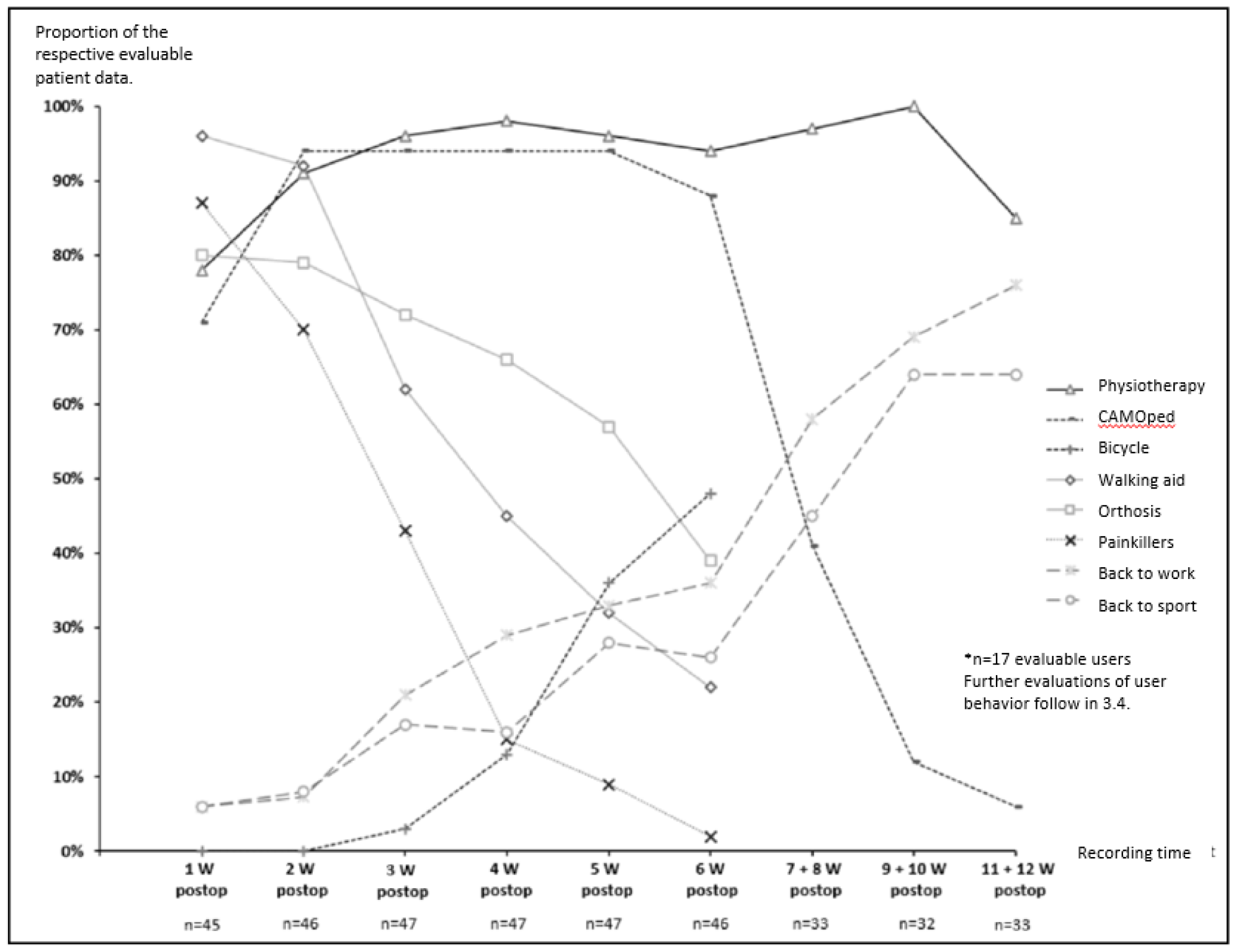
Utilisation of various supportive measures in the course of rehabilitation and return to work and sport in patients after ACLR as part of the CAMOPED study Partial data set; abbreviations: postop, post surgery; W, week.

**Illustration 11.**
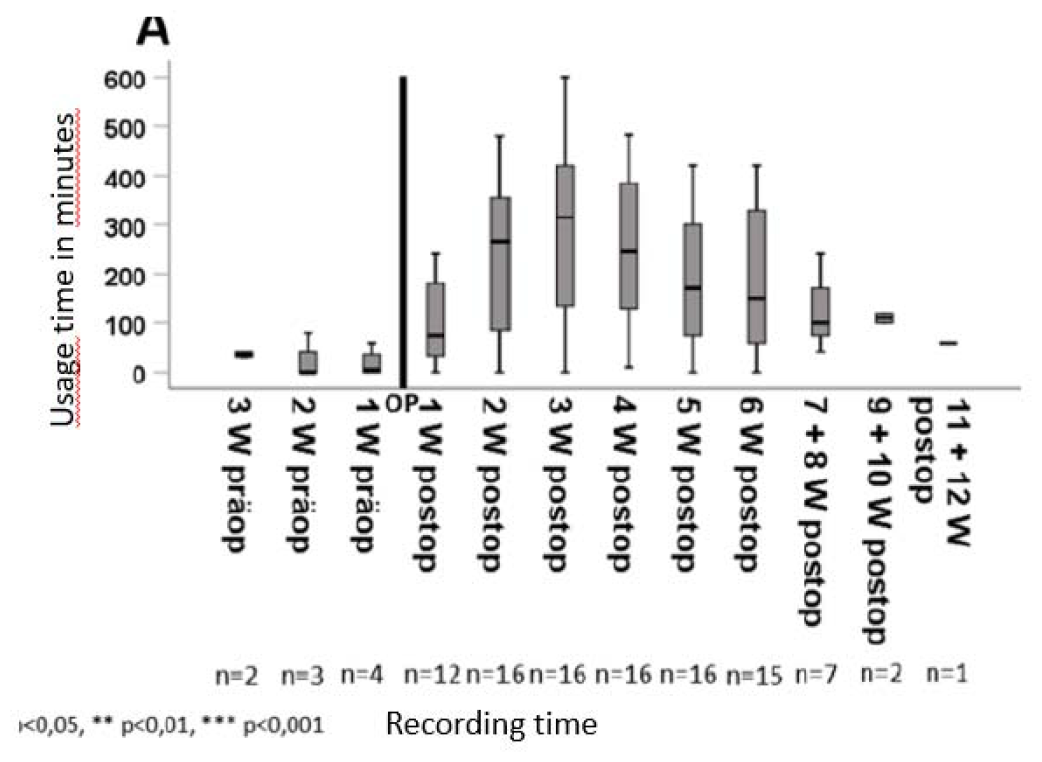
Weekly subjectively reported CAM utilisation times in the course of rehabilitation for patients after ACLR in the CAMOPED study Partial data set; abbreviations: preop, preoperative; postop, post surgery W, week.

The use of physiotherapy and other measures to support rehabilitation (painkillers, walking aid, orthosis, bicycle) was recorded weekly (up to week 6 post surgery) and fortnightly (up to 12 weeks post surgery).

From week 2 to week 10 post surgery, almost all patients received physiotherapy (Illustration 10). The patients who received a CAM splint according to the protocol and whose usage data could be analysed (n=17) used the splint from week 2 post surgery at the latest.

The CAM splint was used for a median of 6 [5-7] weeks on 4.6 [2.9-5.8] days per week. It was used 1.8 [1.1-2.5] times a day for 20 [16.5-26.1] minutes

From week 7, 40% of patients were still using the CAM splint. The proportion fell to less than 10% during the further course of rehabilitation until post surgery week 12.

The intake of painkillers decreased continuously in the first weeks after the operation.

Immediately post surgery, almost all patients used walking aids. Four weeks post surgery, these were only required by less than 50% of patients.

The proportion of patients who cycled increased from week 3 after surgery and by week 6 post-operatively almost 50% of patients were using a bike.

From the 3rd post surgery week, the proportion of patients who returned to work and sport increased. Six weeks post surgery, around a third of patients stated that they had returned to work and around a quarter had started exercising. Twelve weeks post surgery, more than two-thirds of patients were back at work and able to participate in sports activities.

#### 18.3. Predefined subgroup analysis of ACLR with concomitant injuries

In the subgroup of ACLR with concomitant injuries, n=5 were in the IG and n=12 in the KG, so that a further analysis does not appear meaningful.

#### 18.4. Validation of the modified subjective IKCD (LERAS-13)

The results were already presented in 2023 at the congress of the Society for Orthopaedic Traumatological Sports Medicine (GOTS). Below is the German translation of the abstract:

Research question: There is currently no validated measurement instrument for research and clinical use that can be used in the early rehabilitation process after surgical treatment of lower extremity injuries in competitive and amateur athletes. Therefore, a new questionnaire (Lower Extremity Early Rehabilitation Score-13, LERAS-13) was developed and validated to measure return to work, sport and everyday activities in the early and mid-rehabilitation period.

Methodology: The questionnaire consists of questions on the work situation and workloads, return to work and sport as well as 13 items with subjective self-assessments on activities of daily living, from which a total score is calculated. It contains elements from validated knee joint-related function scores and was supplemented with relevant activities from daily life (e.g. crawling, operating foot pedals, walking on rough terrain, pushing or pulling). The questionnaire was developed using the Delphi method and validated according to the COSMIN guideline in German as part of a randomised controlled, single-blind clinical trial (DRKS00021739). At the time of analysis, data from 92 patients (m:w 58:41, age 18-63 years) after rupture and replacement of the anterior cruciate ligament were available up to 6 weeks post surgery. The correlations of the LERAS-13 with validated standard questionnaires from the field (sIKDC, ACL-RSI) as well as the internal consistency (Cronbach’s alpha) and the test-retest reliability were calculated.

Results: The measurement instrument showed very good internal consistency (between 0.87-0.97) and good test-retest reliability (between 0.75-0.92) for all measurement time points. The LERAS-13 correlated with knee joint function (sIKDC, r = 0.5-0.7, each p < 0.001) three and six weeks post surgery. In contrast, there were only minor non-significant correlations with an assessment of the psychological aspects of rehabilitation (ACL-RSI), which demonstrates the high validity of the instrument.

Conclusions: This new, easy-to-use, digital questionnaire is a valid instrument for recording patient-relevant endpoints, in particular return to daily life and work after surgical treatment of lower limb injuries. It closes the gap in existing measurement instruments and enables standardised quantification of individual rehabilitation progress in the early rehabilitation phase for the first time. This means that treatment strategies can be individually evaluated at an early stage and adapted if necessary.

For the validation of the LERAS-13, data from 92 patients were compared with those in Illustration 12 were analysed with the patient characteristics shown in Figure 12.

**Illustration 12.**
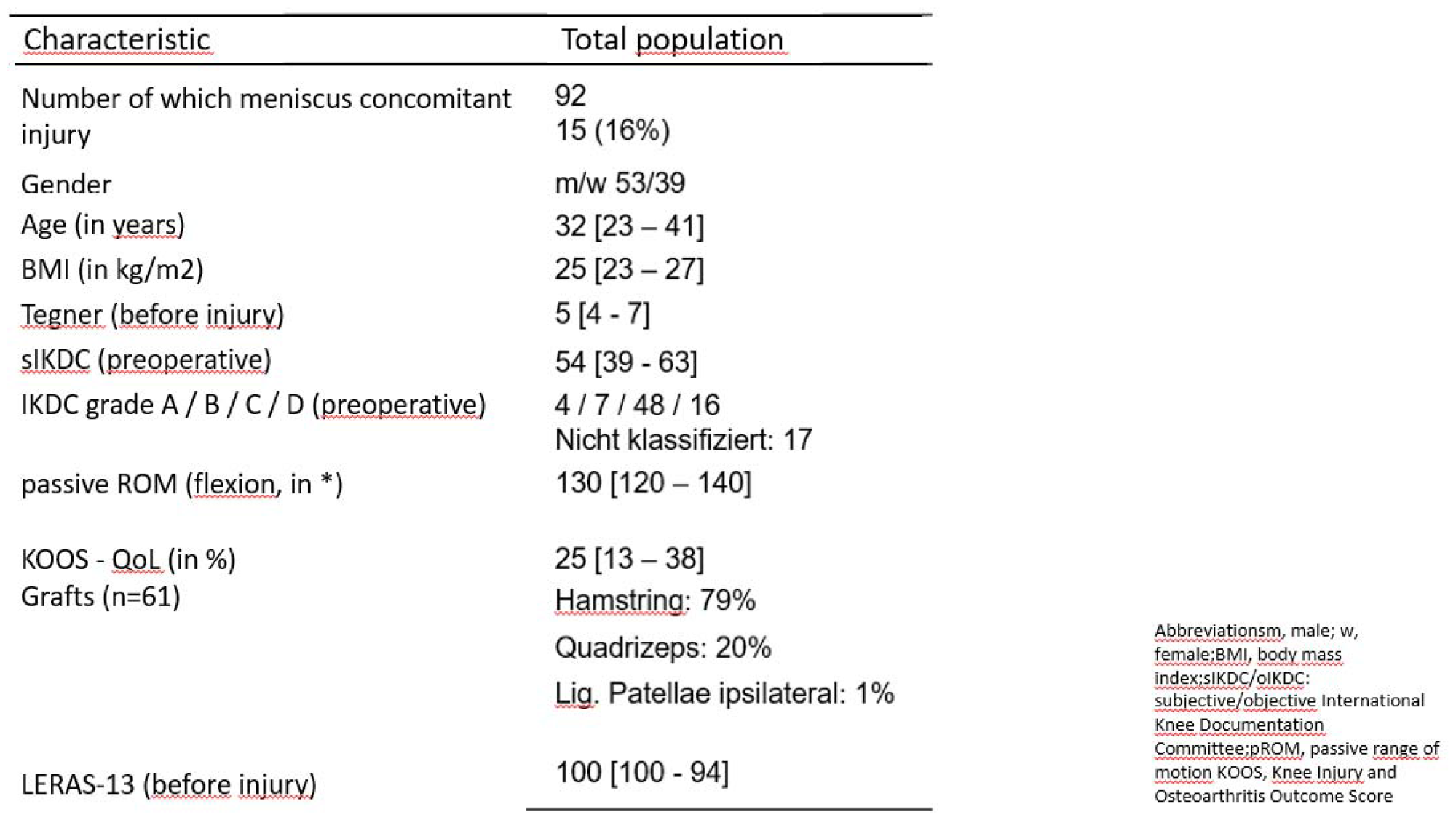
Patient characteristics at study inclusion

The majority of patients had returned to work 7 weeks post surgery and had returned to sport 8 weeks post-surgical.

**Illustration 13.**
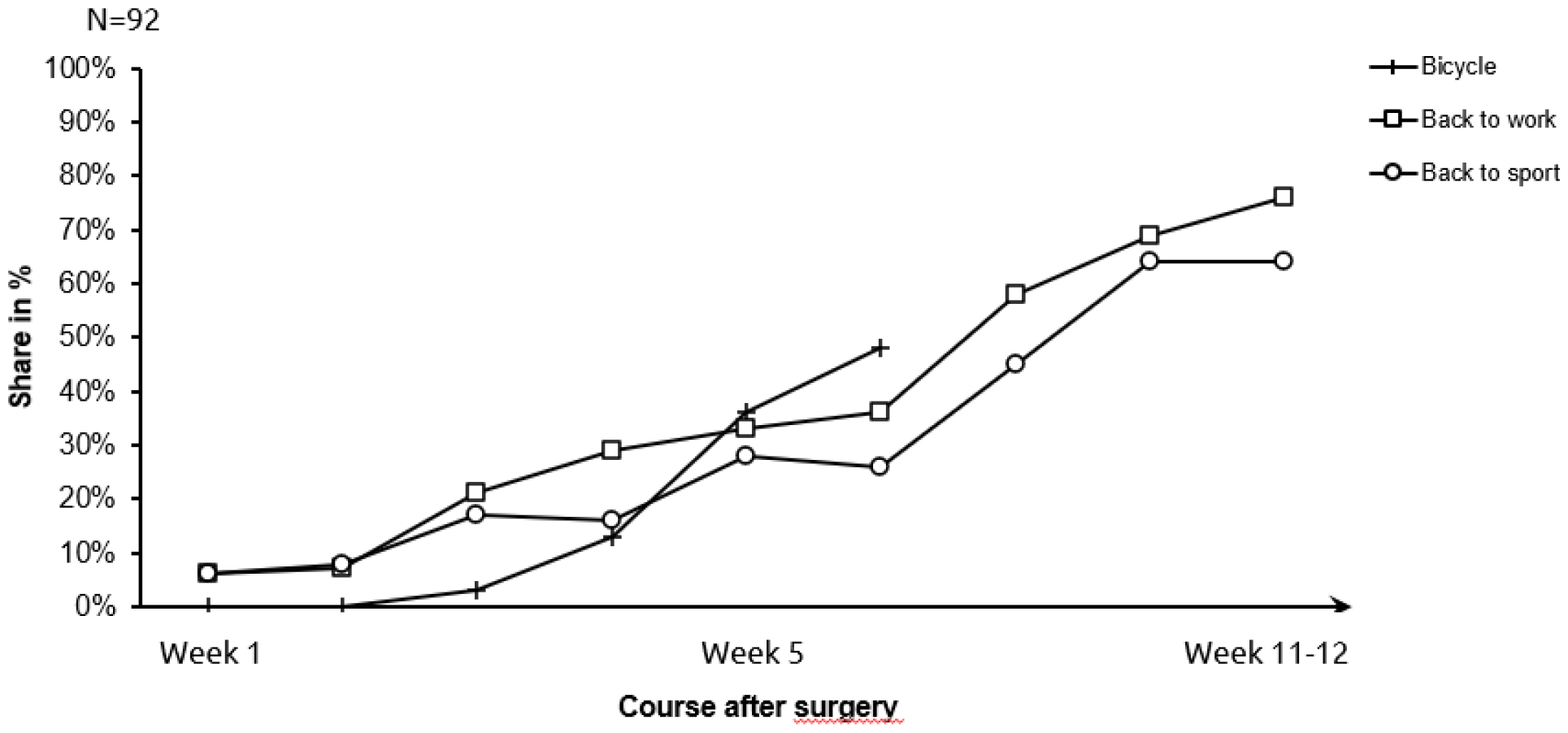
Proportion of patients with resumption of work and sport in the post-surgical course

The LERAS-13 is sensitive to the weekly change during the early rehabilitation phase.

**Illustration 14.**
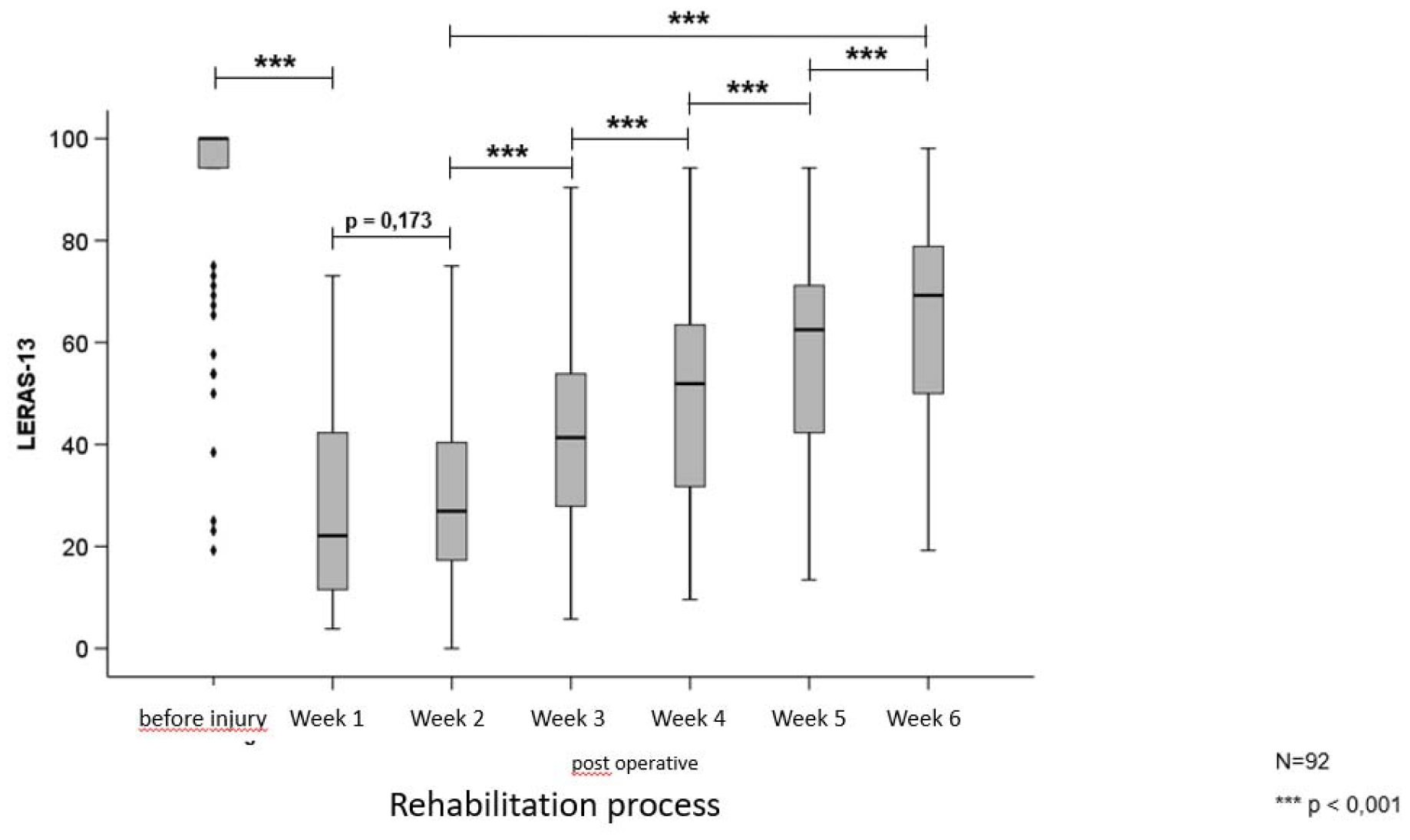
LERAS-13 in the post-surgical course

LERAS-13 differentiated according to the severity of the injury. Patients with isolated ACLR showed a significantly greater increase in LERAS-13 in the early course of rehabilitation than patients with ACLR and meniscus injury (p=0.006).

**Illustration 15.**
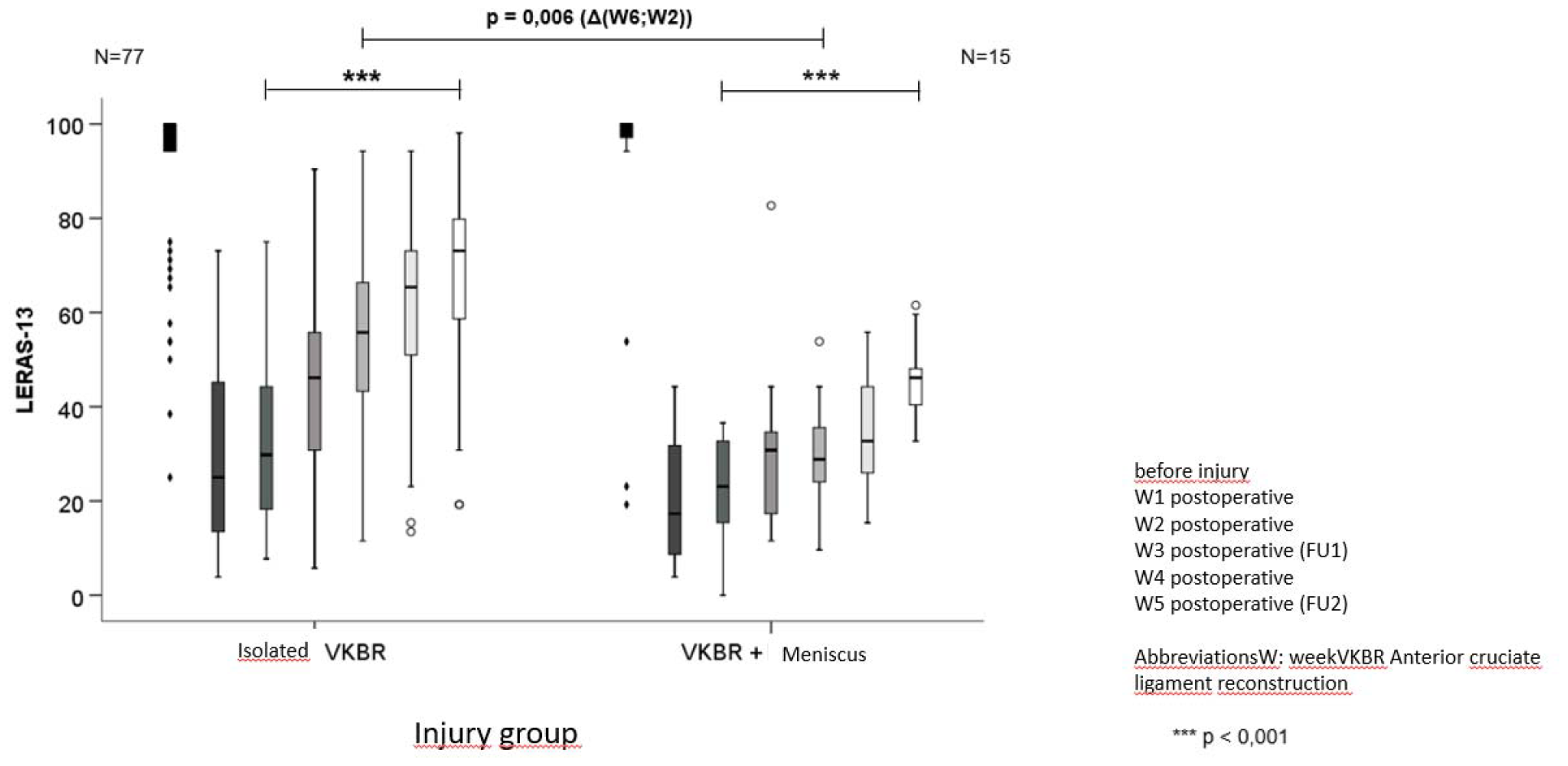
LERAS-13 in the post-surgical course in patients with and without meniscus injury

LERAS-13 depicts the rehabilitation process in an age-specific manner. Patients < 30 years showed a significantly greater increase in LERAS-13 over the course of rehabilitation than patients >= 30 years (p=0.003).

**Illustration 16.**
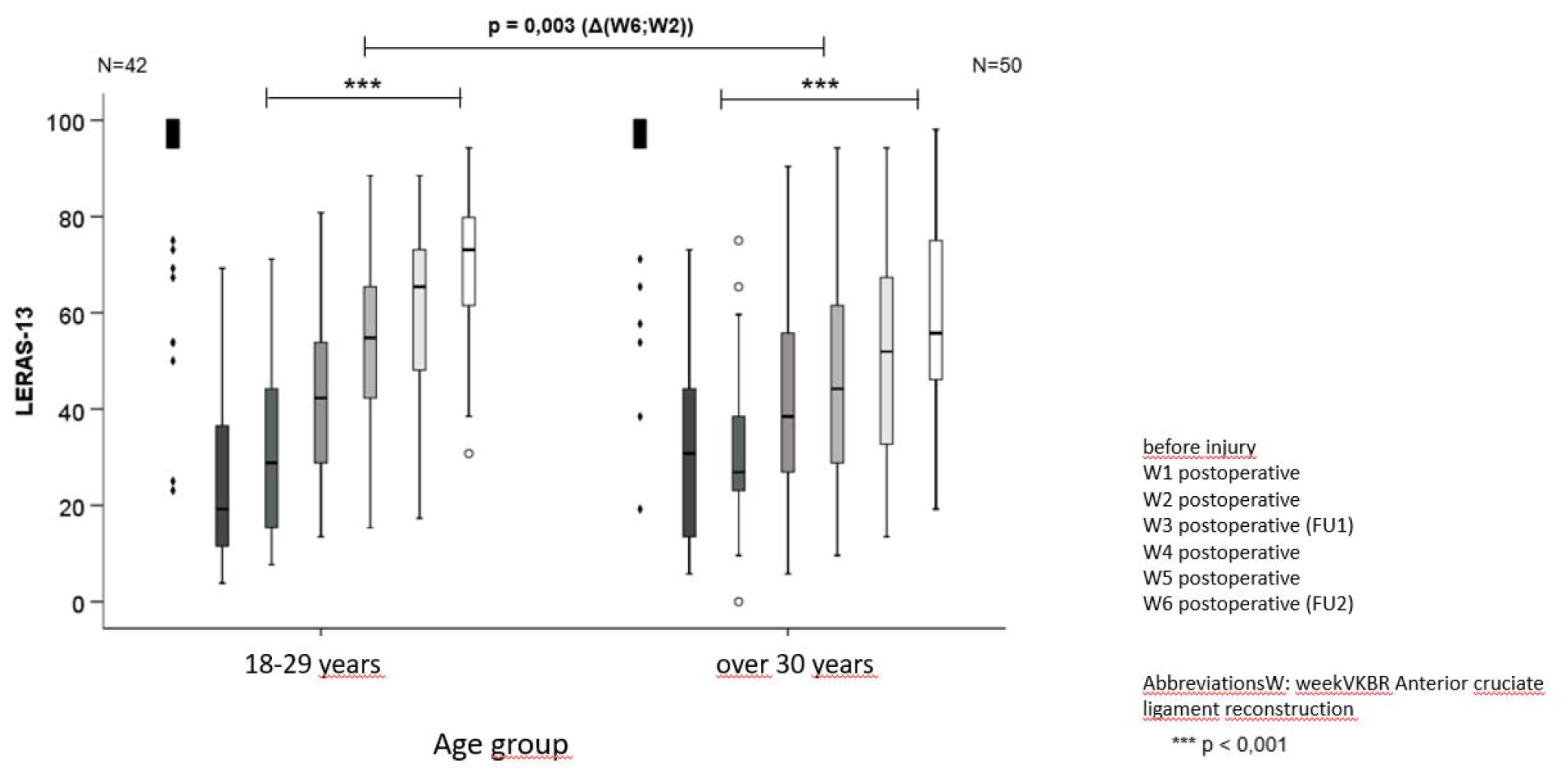
LERAS-13 in the post-surgical course in patients younger and older (>=) 30 years

The LERAS-13 shows good external validity in relation to knee joint function in the early rehabilitation phase.

**Illustration 17.**
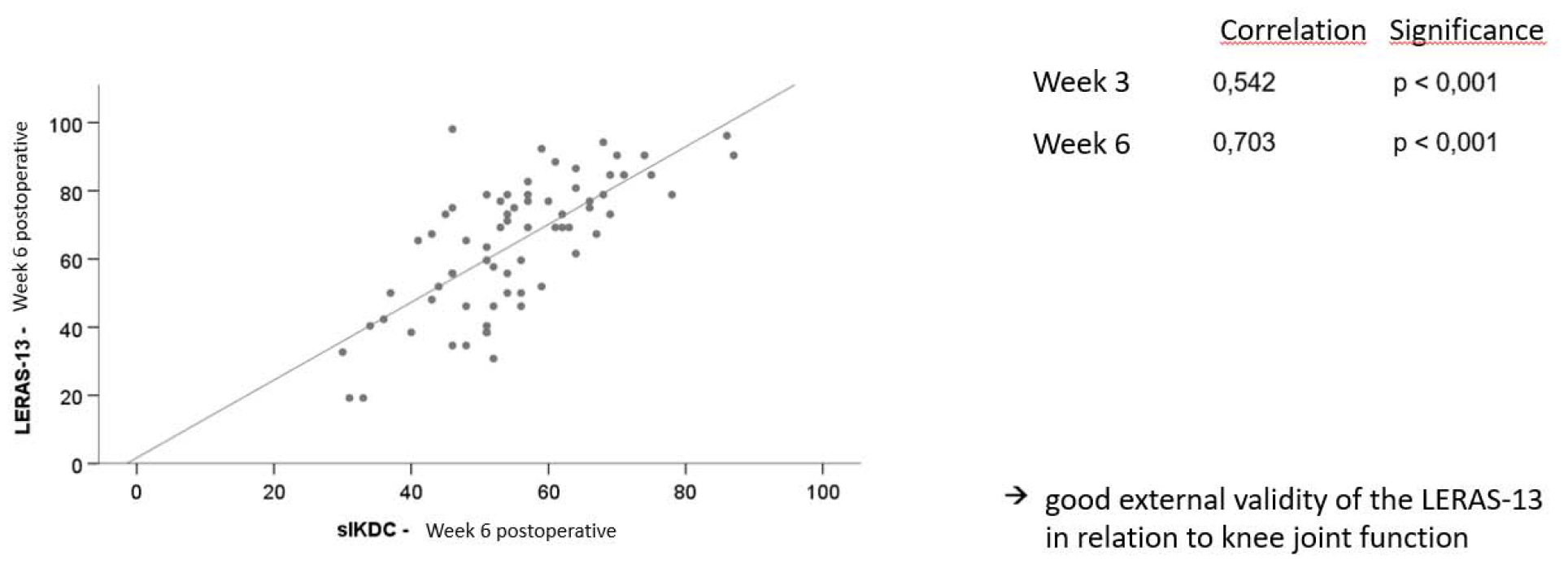
Correlation of the LERAS-13 with the sIKDC in the early rehabilitation phase

The internal consistency and test-retest reliability show a very good measurement accuracy (Cronbach’s alpha 0.92) and a low influence of measurement errors (week 1-6, r = 0.75 - 0.92, p < 0.01).

**Illustration 18.**
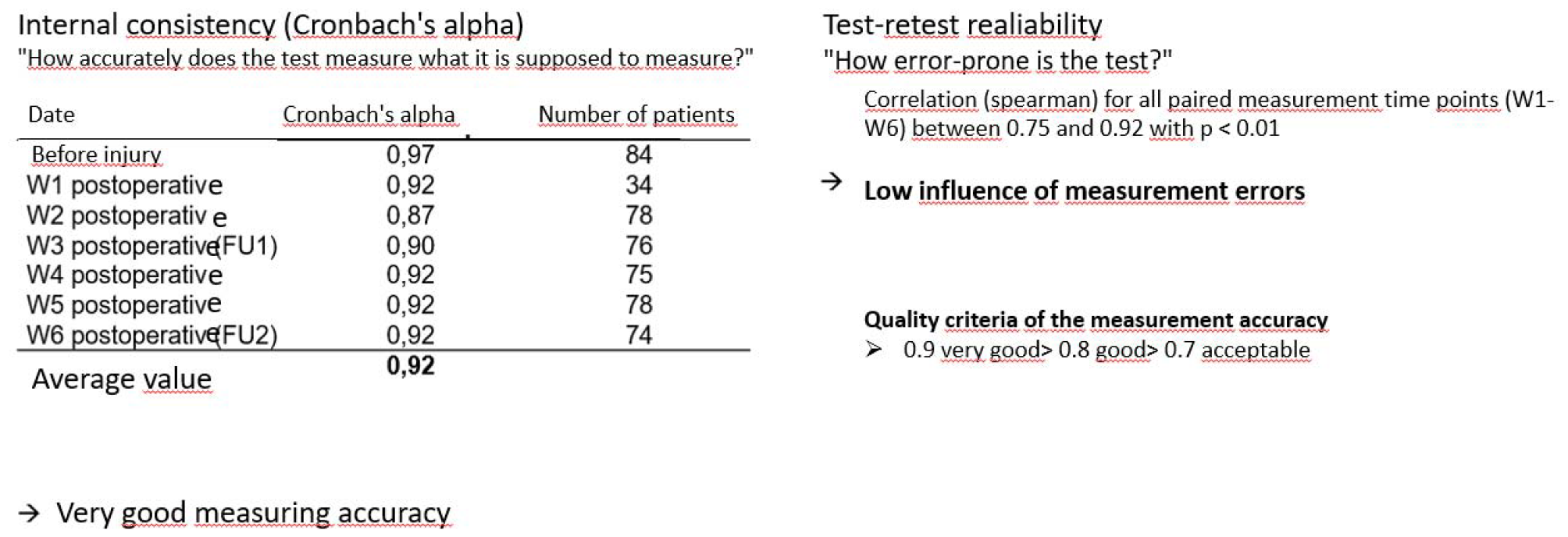
Internal consistency and test-retest reliability of the LERAS-13

The LERAS-13 is a suitable measuring instrument for recording knee function in everyday life (see Illustration 19).

**Illustration 19.**
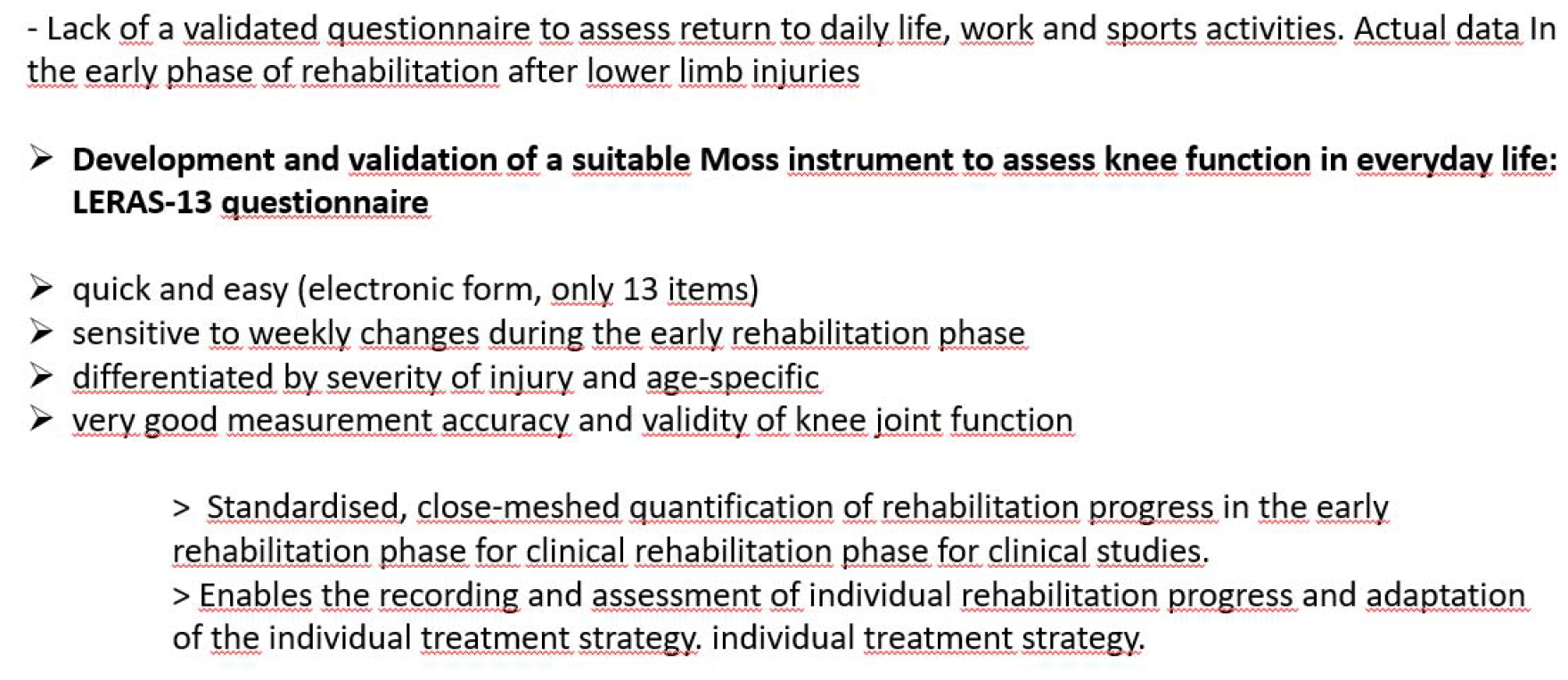
Suitability of the LERAS-13 measuring instrument for recording knee function in everyday life

#### 18.5 Economic efficiency of the CAMOPED

Due to the dropout, this analysis did not appear to make sense.

#### 18.6 Comparability of different surgical procedures (transplant)

Due to the dropout, this analysis did not appear to make sense.

#### 18.7 Superiority of sonographic findings compared to palpation

The results were already published in Int Orthop in 2023 (DOI: 10.1007/s00264-023-05697-x).^28^ Below is the abstract:

Purpose: We sought to externally validate ultrasonography (US) for quantification of suprapatellar effusion size to improve diagnosis and individualised rehabilitation strategies in knee rehabilitation after anterior cruciate ligament reconstruction (ACLR) surgery.

Methods: As part of the ongoing CAMOPED study, 35 patients underwent a US examination. Data were collected at ACLR and after surgery at defined intervals up to one year after surgery. Palpatory assessment was evaluated using the international knee documentation committee (oIKDC).

Results: A total of 164 sonographies showed a strong correlation between palpatory and US effusion (r = 0.83, p < 0.01) with lower deviations of US quantification compared to palpatory quantification Y = 1.15 + 0.15* x. Threshold values for the detection of effusions by palpation and for the differentiation between mild and moderate/severe effusions could be determined (effusion depth: 2.6 mm and 5.8 mm, respectively).

Conclusions: As demonstrated in this multicentre study, the size of suprapatellar effusions can be quantified easily and with high accuracy using standardised bedside ultrasound. Especially for moderate to severe effusions, ultrasound is a practical and reliable tool for outcome measurement that is superior to palpatory assessment with the aim of optimising individual recommendations during the rehabilitation course. In addition, it was possible for the first time to define sonographic thresholds for the detection of an effusion and the differentiation between mild and moderate/severe effusion based on palpation.

### 19. Safety / Adverse events

Adverse events were categorised as AEs and SAEs and assessed for a possible causal relationship with the investigational product.

A total of 90 adverse events were documented, 8 of which were classified as SAEs (Table 11). Two of these events were re-ruptures of the anterior cruciate ligament. The SAEs included one post-surgical syncope, one surgical treatment due to a diagnosis of cervical cancer and four events with scarring or cyclops syndrome requiring surgical resection. In one case, infrapatellar ramus syndrome also occurred (Table 12). None of the events were related to the test product.

**Table 11.** List of adverse and serious adverse events and potential relation to the investigational product.

| **Security**<br>(according to ISO 14155) | **Total** | **Connection with**<br>**CAMOped** |
| --- | --- | --- |
| Incidence | 0 | 0 |
| Adverse events | 91 | 0 |
| of which serious adverse events | 8 | 0 |
| of which re-rupture | 2 | 0 |

**Table 12.** List of serious adverse events and potential relation to the investigational product.

| **Event** | **Period of occurrence (in**<br>**post surgery**<br>**weeks/months)** | **Causal relationship to**<br>**the CAM splint** | **Intervention** |
| --- | --- | --- | --- |
| Post surgery syncope | 1 week | no | Stationary monitoring,<br>exclusion of LAE |
| Surgical treatment for<br>uterine cervical cancer | 8 weeks/9 weeks | no | None |
| Re-arthroscopy and scar<br>release for joint stiffness | 6 months | no | Re-arthroscopy and<br>arthrolysis |
| Cyclops syndrome | 12 months | no | Re-arthroscopy with<br>conglomerate removal |
| Cyclops syndrome | 7 months | no | surgical resection,<br>arthrolysis |
| Ramus infrapatellaris<br>syndrome, cyclops<br>syndrome | 14 months | no | Surgical neurolysis and<br>arthrolysis |
| Re-rupture of the ACL | 11 months | no | Revision |
| new distortion trauma<br>with re-rupture of the<br>ACL | 12 months | no | Revision |

## IV Summarising assessment / discussion

### 20. Limitations

The main limitation of this study results from the premature discontinuation of the study due to the ethically unacceptable continuation of the study with a control group without CPM splint for all patients, as decided by the LKP together with the principal investigators after discussion of the interim report (report interim analysis V0.7 20220919) and communicated to the G-BA on 23 January 2023 (see Appendix 4).

The reason for this was the use of “Continuous Passive Motion” (CMP) splints as part of the standard therapy in the rehabilitation phase following treatment of ruptures of the anterior cruciate ligament. However, the control group in the CAMOPED study was not allowed to receive a splint during the rehabilitation phase in accordance with the protocol. At the time (11/2022), this no longer corresponded to the current reality of care. As the study data showed at the time, there has been a high drop-out rate (28%) of patients since December 2021 because they demanded the standard therapy in the control group or the control group should be provided with the standard therapy. The LKP and the principal investigators understood that sports bans in 2020 to 2021 reduced the eligible injuries to such an extent that the study could not be completed as planned. The group of peers considered the continuation of the study with the current design to be outdated and unethical.

The primary hypothesis, according to which one of the treatments leads to a significantly greater improvement in the subjectively assessed knee joint function within the first 6 weeks of the post surgery rehabilitation phase in a group comparison, can therefore not be answered.

In addition, the influence of the significantly higher value of the sIKDC post surgery compared to the CG on the only half as high proportion of additional meniscus interventions with the resulting lower post surgery restriction of knee joint function in the group is responsible for the absolute difference in the course to FU2. The sIKDC values for FU2 did not differ between the two treatment groups. This effect is most likely due to the relatively small number of cases in the treatment groups and their subgroups. These results could be analysed for the first time after the complete data sets from the 5 institutions were available.

The sIKDC score was recorded as a patient-relevant endpoint in the CAMOPED study. The use of the sIKDC score in the early phase of rehabilitation is seen as the main obstacle to a positive result in this study, as the score is not suitable for the early rehabilitation phase and may not be sensitive enough to capture the actual patient-relevant changes. This was already discussed before the start of the study and at the same time various new measurement variables were therefore validated for this question to record patient-relevant endpoints: these are the sonographic quantification of knee joint effusion and a score for assessing knee joint function in the early and middle rehabilitation phase (LERAS-13). The respective results were presented and discussed at specialist congresses over the last two years and have now been accepted for publication in a high-ranking journal (for results, see also chapter 18.4 and 18.7).

The G-BA’s timetable envisaged the final report of the study in autumn 2023. As individual centres continued to include patients in the study until the LKP’s written notification to the G-BA, not all long-term examinations are available here at the 12-month point. However, as these data were classified as relevant from a scientific point of view, the study will not be completed until the last visit of the last patient has been completed, as recommended by the LKP.

### 21. Generalisability of the results

#### 21.1 Target population and progression

As shown in the demographic data, the target population was mapped in this study in terms of gender and age distribution and the activity index. The study design also included patients with strongly divergent physical constitutions and physical activity levels, in line with the target population. ^10, 29, 30 31 32^

At study inclusion (T0), the relevant confounding preoperative variables (age and physical activity) were equally distributed between the two study groups. Thus, the randomisation can be considered successful.

The values of the analysed variables for the assessment of knee joint function (subj. IKDC, KOOS QoL, obj. IKDC) were within the expected ranges at inclusion and throughout the course of the study ^15 16 17^.

The concomitant injuries, which often only become visible during the operation and which, from a clinical perspective, have the greatest influence on the post-operative rehabilitation process, were to be depicted in the study in accordance with the protocol. In the various centres, the plastics were mostly performed using hamstring tendon reconstruction, while one centre implanted quadriceps tendons. This means that the target population is also represented with regard to this point.

In the overall study population, the sIKDC score decreased post surgery (time point T4) compared to the baseline values recorded at inclusion time point T0 and increased again in the following weeks during the course of rehabilitation. The sIKDC score reached the baseline value 6 weeks post surgery and was higher than the pre-operative value 12 weeks post surgery. Thus, the course of the subjectively assessed knee joint function in the overall study collective corresponds to the expected course of the target population.

Similarly, KOOS Q1-Q4 was used to map knee-related quality of life at inclusion and over the course of the target population.

#### 21.2. Effect of the active motion splint on knee joint function after ACLR intervention

Due to premature termination of the study for ethical reasons, it was not possible to achieve a sufficiently large number of study inclusions that could have been treated in the control group according to protocol as standard therapy without the use of a CPM splint. Therefore, no conclusive statement can be made here.

#### 21.3 Further explorative analyses

The data on the use of CAMOped by patients and the lack of reported problems demonstrate its ease of use in the home. The safety of this study (see chapter 19).

The results obtained in this study for the validation of sonography for the quantification of knee joint effusions and of LERAS-13 for the assessment of early rehabilitation success after ACLR appear to be valid according to internal and external criteria, sufficiently sensitive and suitable for recording a primary endpoint for a future study on this issue.

### 22. Interpretation

For the reasons mentioned above, it is ultimately not possible to assess the possible superiority of the active motion splint in terms of improving knee joint function.

Rather, a future study should:

- A comparison of the CAM splint with the current standard of care, the CPM splint, should be sought. This should logically test for non-inferiority and prove the benefit of the CAM splint.
- The newly developed LERAS-13 score can be used as a measuring instrument to assess the patient-relevant endpoint in the early and middle rehabilitation phase.

The proposal for a non-inferiority test of CAM splints against CPM splints is hereby submitted to the Federal Joint Committee (G-BA).

## V Further information

### 23. Registration

The study has been registered under the number DRKS00021739 prior to enrolment of the first patient.

### 24. Protocol

The complete protocol (test plan code: 1910_44_19) is attached to this report as Annex 1 Test plan RCT CAMOped V1.2 20201214.

### 25. Financing

The project was financed 100% by the manufacturer, OPED GmbH. The sponsor has no share in the concept, implementation, interpretation, evaluation or reporting.

## Data Availability

All data produced in the present study are available upon reasonable request to the authors

## Appendices can be issued by the publisher in German on request

Table of contents of the annexes:

- Appendix 1 Test plan RCT CAMOped V1.2 20201214
- Appendix 2 SAP CAMOped V1.1 20201214
- Appendix 3 Report Interim Analysis V0.7 20220919
- Appendix 4 Recommendation LKP_CAMOPED21012023
- Annex 5 4th reply G-BA 20201214

## List of abbreviations

Abbreviation: Meaning
ACL: anterior cruciate ligament
AC: Adverse event
ADE: Undesirable effects of the product (adverse device effect)
CAM: Continuous active motion (active motion rail)
CPM: Continuous passive motion (passive motion rail)
DD: Product defect (device defect)
EC: Ethics Committee
FU: Follow-up (follow-up examination)
G-BA: Joint Federal Committee
GAS: Goal Attainment Scale
IG: Intervention group
IQWiG: Institute for Quality and Efficiency in Health Care
CG: Control group
KOOS: Knee Injury and Osteoarthritis Outcome Score
LERAS-13: Lower Extremity Early Rehabilitation Score-13
LKP: Head of the clinical trial (principal investigator)
LCL: Lateral collateral ligament
MCID: Minimal clinically important difference
MCL: medial collateral ligament
PROM: Passive range of motion
RCT: randomised controlled trial (randomised controlled study)
SADE: Serious undesirable effects of the product (serious adverse device effect)
SAE: Serious adverse event
SCB: Substantial clinical benefit
IKDC: International Knee Documentation committee
TMF: Trial Master File
ACL: Anterior cruciate ligament
ACLR: Anterior cruciate ligament rupture

